# Public Opinion and Sentiment Before and at the Beginning of COVID-19 Vaccinations in Japan: Twitter Analysis

**DOI:** 10.1101/2021.07.19.21260735

**Authors:** Qian Niu, Junyu Liu, Masaya Kato, Yuki Shinohara, Natsuki Matsumura, Tomoki Aoyama, Momoko Nagai-Tanima

## Abstract

**Background:** The pandemic of COVID-19 is causing a crisis in public health, food systems, and employment. Vaccination is considered as one of the most effective ways for containing the pandemic, but widespread vaccine hesitation on social media may curtail uptake progress. Fully comprehending public sentiment towards the COVID-19 vaccine is critical to building confidence on the vaccines and achieving herd immunity, especially in Japan with inadequate vaccine confidence.

**Objective:** This study aims to explore the opinion and sentiment towards the COVID-19 vaccine in Japanese tweets, before and at the beginning of large-scale vaccinations.

**Methods:** We collected 144,101 Japanese tweets containing COVID-19 vaccine-related keywords between August 1, 2020, and June 30, 2021. We visualized the trend of number of tweets and identified the critical events that triggered a surge and provided high-frequency unigram and bigram tokens. Also, we performed sentiment analysis and calculated the correlation of number of tweets and positive/negative sentiments with infection, death, and vaccinated cases. we also used the latent Dirichlet allocation (LDA) model to identify topics of tweets. In addition, we conducted analysis on three vaccine brands (Pfizer/Moderna/AstraZeneca).

**Results:** Daily number of tweets continued growing and the growth accelerated since the large-scale vaccinations in Japan. The sentiment of around 85% tweets were neutral, and the negative sentiment overwhelmed the positive sentiment in the other tweets. Number of tweets strongly correlated (r≥0.5) with infection/death/vaccinated cases, and the number of negative tweets correlated strongly with the number of infection/death cases but weakened after the first vaccination in Japan. LDA identified three public-concerned topics: vaccine appointment and distribution strategy; Different vaccines development progress and approval status of countries; Side effects and effectiveness against mutated viruses. Among vaccines of the three manufactures, Pfizer won the most attention and Moderna the least.

**Conclusions:** Our findings indicated that negative sentiment towards vaccines dominated than positive sentiment in Japan. Changes in number of tweets and sentiments might be driven by critical events related to the COVID-19 vaccine, and negative sentiment continued increasing when numerous adverse accidents occurred at the beginning of large-scale vaccinations. Under the negative sentiment, the concerns of three vaccine brands remains effectiveness and safety with slight differences. The policymakers should provide more evidence about the effectiveness and safety of vaccines and optimize the process of vaccinations.

## Introduction

A novel coronavirus causing COVID-19 was first identified in December 2019 [1]. Until June 30, 2021, the cumulative confirmed and death cases were approximately 181 million and 3.9 million globally [2], of which 798,159 confirmed cases and 14,740 death cases were reported in Japan [3]. The pandemic has had a significant adverse effect on individuals, governments, and the global economy [4].

Without effective treatments and medications, herd immunity by vaccination is considered as one of the most effective ways for containing the pandemic. Global efforts to develop and test COVID-19 vaccines have intensified since the declaration of pandemic in March 2020 [5]. Several vaccines have been developed and authorized in record time. The Pfizer/BioNTech vaccine was authorized in the UK on 2 December 2020. Japan lagged other developed countries, licensing Pfizer on February 14, 2021, and Moderna and AstraZeneca on May 21, 2021. Japan experienced significant delays in COVID-19 vaccinations [6], with a vaccination rate of less than 4% until May 21, 2021. Meanwhile, Tokyo is preparing to host the 2020 Summer Olympics on July 23, 2021, yet only 13.6% of the population was fully vaccinated as of June 30. To reach herd immunity, at least 70% of the population needs to be vaccinated [7], but high vaccination rate relies heavily on the public willingness of vaccinations. Vaccine hesitancy is one of the ten main risks to world health according to the World Health Organization (WHO) [8,9]. Japan ranked among the countries with the lowest vaccine confidence in the world, which might result from the crisis of confidence towards the human papillomavirus (HPV) vaccine in 2013 [10]. The reluctance to be vaccinated against COVID-19 is particularly worrisome from a public health standpoint.

It is critical to understand reasons for vaccine hesitancy in Japan. Surveys are often used to gather data on vaccine hesitancy, but it is often costly and time-consuming, and only inflects relatively short-term situations and limited samples. In contrast, social media research can be cheaper, and are more practical for conducting almost instantaneous information of the opinions and sentiments of large population. There were approximately 90.6 million Japanese social media in 2020, accounting for 72.48% of the total population [11]. Besides, the ongoing COVID-19 pandemic led to increasing use in social media as a venue for discussion of vaccine-related issues [12], and opinions of social media users may affect individual opinions and results in vaccine hesitation or refusal [13]. Notably, misinformation on social medias can undermine public confidence in science and public health authorities, intensifying the vaccine hesitancy [14,15]. Addressing vaccine hesitancy and helping to boost public trust in vaccinations by investigating public attitudes towards vaccinations may lead to greater vaccination coverage [16].

During the pandemic, research widely applied social media to mine opinions and sentiments towards the COVID-19 vaccine in various countries, but Japan is still in a gap. Several studies have linked social media activity to vaccine hesitancy and anti-vaccine campaigns [17–20]. Tangherlini and colleagues [17] analyzed vaccine hesitancy drivers on blogs and reported that parents may utilize these forums to spread vaccine opposition views to other parents. Bonnevie and colleagues [14] studied the evolution of vaccine resistance in the US by analyzing Twitter discussion topics and found that prominent Twitter accounts were responsible for a significant percentage of vaccine-opposition messages. Cossard and colleagues investigated the extent to which vaccination discussions may influence vaccine hesitancy, as well as how skeptics and proponents’ networks are structured differently [21]. Their results demonstrated that anti-vaccine community clusters are more united than pro-vaccine community clusters. Lyu and colleagues proposed that public discussion is driven by major events, and vaccine sentiment around COVID-19 is increasingly positive in six countries, showing higher acceptance than previous vaccines [22]. The findings of several studies indicate positive attitudes in Australia, the UK, and the US, but more positivity is needed to boost vaccination rates to attain herd immunity [23,24]. Marcec and colleagues checked the sentiment towards different vaccine brands and indicated the sentiment in some countries towards the AstraZeneca/Oxford vaccine seems to be decreasing over time, when sentiment regarding Pfizer and Moderna vaccines kept positive and stable [25]. Samira and colleagues took extra effort to conduct large-scale research on people’s views on vaccines by Twitter [26]. Japan has over 50 million Twitter users, constitutes approximately 45% of the total population [27], providing a powerful data source for large-scale opinion and sentiment mining.

This study aimed to examine the public opinion and sentiment expressed by Japanese Twitter users before and at the beginning of the large-scale COVID-19 vaccinations between August 1, 2020, and June 30, 2021. We summarized the trend of number of tweets and extracted topics from vaccine-related tweets. We also calculated the correlation between the number of infection/death/vaccinated cases and the total number of tweets and number of tweets of different sentiments over time. We further analyzed the different opinions and sentiments towards three manufacturers, Pfizer, Moderna, and AstraZeneca. To our knowledge, our study is the first application of sentiment analysis related to public health via social media in Japan. Findings from this research can help governments, policymakers, and public health officials understand factors that motivate and cause hesitance in the public towards vaccinations and provide evidence for planning, modifying, and implementing the tailored vaccine promotion strategy to achieve herd immunity.

## Method

### Data Extraction and Preprocessing

A large-scale COVID-19 Twitter chatter dataset collected and maintained by Georgia State University’s Panacea Lab [28] was used in this study. The tweet IDs, posting date and time, and the languages of all tweets are provided in the dataset. In this study, 1,305,308 Japanese tweets from August 1, 2020, to June 30, 2021, were downloaded. We first extracted the IDs of Japanese tweets (some are mixtures of Japanese and English) and then collected the tweets using the Python package Tweepy. All the English words in the tweets were transferred from full width to half-width forms, and lowercased. The tweets were filtered by keywords (Appendix 2, table 1) related to vaccinations. After data cleaning, 144,101 tweets with selected keywords were used for further analysis. Official data on the number of deaths, infections, and vaccinated cases were collected from the website of the Ministry of Health, Labor and Welfare [29], the Prime Minister’s Office [30], and the Cabinet Secretariat [31] of Japan.

**Table 1.** The correlation of the positive/negative sentiment with the daily infection, death, and vaccinated cases. †

| Sentiment | Example |
| --- | --- |
| Positive | First vaccine! Muscle injection, surprisingly not painful. |
| Negative | I plan to be vaccinated myself, but in the absence of medium- and long-term |
| Neutral | Five vaccination sites are available for reservation in Higashinari Ward. |
| Mixed | I heard that the Ministry of Education, Culture, Sports, Science and Technology will issue vaccination certificates for students planning to study abroad. That's great, but isn't the Ministry of Health, Labor and Welfare supposed to do the vaccination |

† The tweets are paraphrased to protect users' privacy.

### Unigram and Bigram Token Analysis

Tokenization is a fundamental step in many natural language processing (NLP) methods, especially for languages like Japanese, which do not have spaces between words. We tokenized all tweets and analyzed the unigram and bigram tokens. The website links, special characters, numbers, and “amp” (ampersands) were removed from the tweets before tokenization. Then, the Python packages SpaCy and GiNZA were used to remove the Japanese and English stop words and implement word segmentation. The Python package scikit-learn was used to convert the segmented sentences into unigram and bigram tokens and to calculate the counts of tokens. The plots of the counts of unigrams and bigrams are shown in Appendix 1, Figure 1-3. We then built word networks on unigram tokens and bigram tokens separately. For unigram tokens, the top 200 tokens were selected. The cosine similarity (>0.3) [32]between each pair of selected tokens was calculated. A word network was created with all the selected tokens as vertices and the scores as the weights of the edges. For bigram tokens, the top 100 tokens were selected. A word network was created with all the words in the selected tokens as vertices, and with the count of tokens as the weights of the edges between the words.

**Figure 1.**
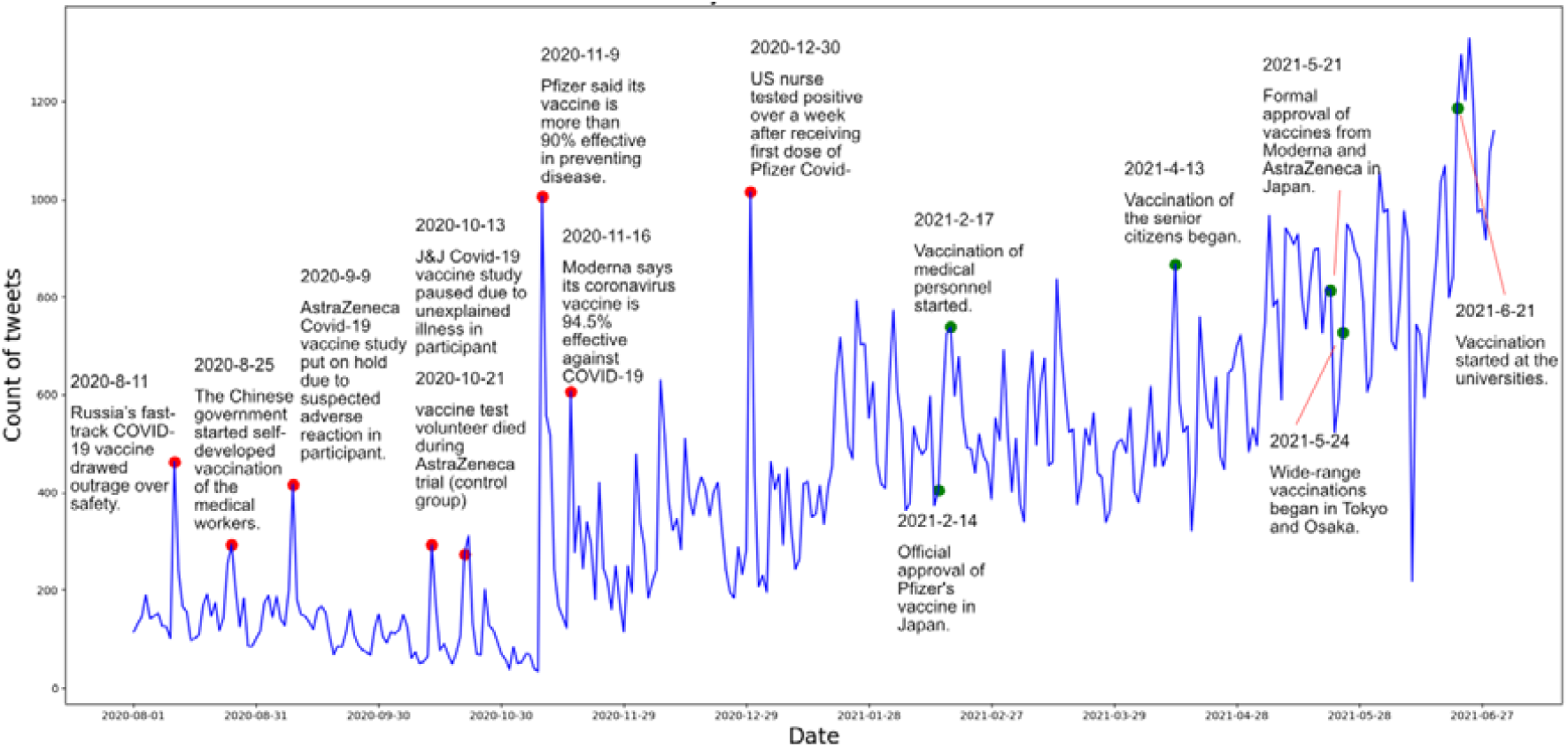
Trends of vaccine-related tweets with key events collected between August 1, 2020, and June 30, 2021. The red points are detected peaks and green points are important events related to vaccinations in Japan.

### Latent Dirichlet Allocation

LDA [33] is an unsupervised generative probabilistic model widely used in topic modeling. We applied LDA modeling of vaccine-related tweets in Japan for easier comparison with recent studies of similar themes in other countries using LDA [22,24]. To apply the LDA method, we first generated the document-term matrix, which recorded the token frequencies in each tweet. All the tweets were put into a list, where each tweet was made into unigrams and reconnected by spaces between neighboring tokens. A document-term matrix was generated on the reconnected tweets. Similar to [24], we also adopted the R package ldatuning [34] for topic number selection. The score of four different metrics was calculated for the topic numbers from two to 50 (Figure 1 of the Appendix 1). The topic number with lower scores for the metrics of “Arun2010” [35]and “CaoJuan2009” [36], and higher scores for the metrics of “Griffiths2004” [37] and “Deveaud2014” [38], is more suitable for LDA modeling. In this study, the “Deveaud2014” reached the highest score and the rest methods all reached second-best performances on three topics over all the vaccine-related tweets. Therefore, we built three-topic LDA models using the Python package scikit-learn, for corpora including all the tweets, the tweets of different sentiments, and the tweets of different vaccine brands. The results were made into bar plots of different words and word clouds for the convenience of analysis and visualization.

### Sentiment Analysis

Lacking efficient models and labeled corpora for a mixture of Japanese and English, we didn’t propose or fine-tune a model for sentiment analysis. Instead, the Amazon Web Services (AWS) supporting multiple languages was chosen for this task. The sentiment analysis includes four labels: positive, negative, neutral, and mixed. For each tweet, the model predicts the label and provides the score for each label. Some examples of Tweets of different sentiments are in Table 1.

To determine the long-term tendency of the statistical data on public attitudes, we calculated the Pearson correlation coefficient (*r*) between the daily number of positive/negative tweets and the daily statistics of death, inflection, and vaccinated cases using the Python package NumPy. The closer the absolute value of the *r* to 1, the stronger is the linear correlation between X and Y. In this study. We calculated the correlations before and after the start of vaccinations in Japan to determine whether vaccinations had an influence on the correlations. The *r* of two sets of data X and Y is

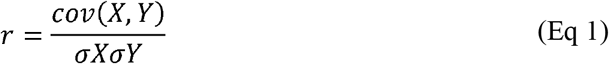

### Peak Detection on Daily Trends

To provide an overview of the data, we plotted the trends of the daily number of total tweets and positive/negative tweets. For more precise analyses, the peaks in the data were labeled on the plots. For a human-like but objective selection, the peaks were determined by an algorithm with reference to the Weber–Fechner law [39] instead of human observation. The peaks of each month were selected using the following ratio:

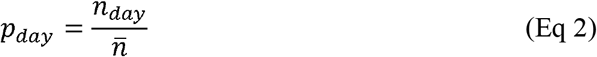

where *n*_*day*_ is the number of tweets for the selected day and 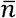 is the average number of daily tweets in the month. If the ratio *p*_*day*_ is higher than a threshold *λ*, the number of tweets of that day is judged as a peak. We then selected the valid peaks from all the peaks. Peaks with less than *n*_*min*_ tweets and peaks within *d* days from the previous peak were abandoned. In this study, *λ* = 1.8, *d* = 5, and *n*_*min*_ *=* 200 for the total number of tweets, and *n*_*min*_ *=* 40 for the number of positive/negative tweets. For all the detected peaks, we checked the tweets and provided the headline vaccine-related news in the tweets that day.

## Results

### Overview of the Data

We counted the number of vaccine-related tweets every day and the trend are in Figure 1. Headline news marking the milestones in Japan’s vaccinations was marked on the curve. The number of daily vaccine-related tweets continually increased over the whole time period. Before November 9, 2020, the number of daily vaccine-related tweets was around or below 200. There are several peaks in the number of related to some important vaccine-related news. On August 11, 2020 (n=463; n indicates the number of tweets), Russia approved the world’s first COVID-19 vaccine [33]. On August 25, 2020 (n=293), the Chinese government initiated the vaccinations of medical workers with self-developed vaccines. On September 9, 2020 (n=416), the AstraZeneca COVID-19 vaccine study was put on hold because of suspected adverse reactions in participants. Two relatively small peaks (n=292, n=274) were related to negative news about clinical trials in October. The first sharp peaks in the number of daily vaccine-related tweets were on November 9, 2020 (n=1006) and November 16, 2020 (n=606): Pfizer stated that its vaccine was 90% effective [35], and Moderna reported the effectiveness of 94.5%. The second surge in the number of daily tweets is December 30, 2020 (n=1015), when a US nurse tested positive over a week after the first dose vaccinations [36]. No additional peaks were detected in the following period, but the daily number of vaccine-related tweets continued increasing, especially after April, 2021. This may be related to large-scale vaccinations in Japan, which is a long-term event across months. On February 14, 2021 (n=405) and May 21, 2021 (n=814), Japan approved the use of Pfizer, Modena, and AstraZeneca vaccines. Japan has adopted a sequential approach to vaccinations by opening appointments on February 17 (n=739), April 13 (n=867), May 24 (n=727), and June 21 (n=1187), 2021, for medical workers, senior citizens over 65 years of age, residents of highly infected areas (Tokyo and Osaka), and university students.

We also created the word networks of unigram and bigram tokens for all vaccine-related tweets. We examined the similarities among top-200 unigram tokens in Figure 2. The token number was selected according to the frequency of each token in the corpus (Figure 2 of the Appendix 1). A major network of words consisted of keywords associated with concerns of vaccines such as “news,” “mutation,” “infection,” “side effect,” “danger,” “senior citizens,” “clinical,” and “prevention.” Another noteworthy sub-network was composed of keywords that were related to countries: “The United Kingdom,” “India,” “Germany,” “China,” “USA,” “Russia,” “Korea,” “Europe,” and “Brazil.”

**Figure 2.**
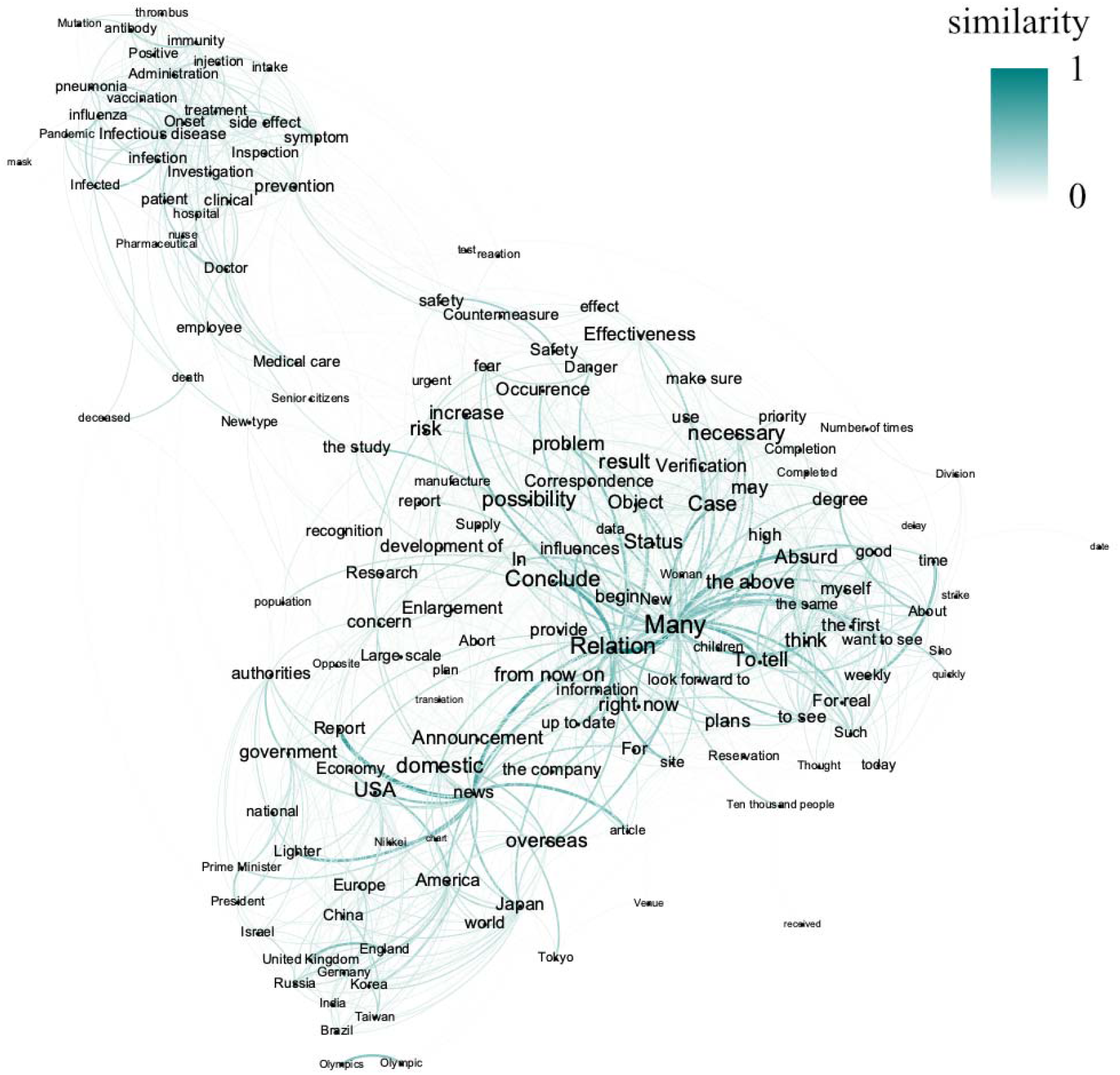
Network of similarity (>0.30) among top-200 unigram tokens (English Version). The edges in the graph are the similarity between unigram tokens, where thicker and more opaque edges indicate stronger similarities.

Figure 3 shows the network of top-100 bigram tokens. The number of tokens in the network was selected according to the frequency of each token in the corpus (Figure 3 of the Appendix 1). According to Figure 3, frequently used words in the tokens were “Japan,” “Astra,” “Medical care,” “Mutation,” “Pfizer,” and “infection.” Moreover, the word “government” was linked to several tokens such as “Tokyo,” “Olympics,” “economy,” and “Prime minister.” There were also a few word pairs apart from the main network.

**Figure 3.**
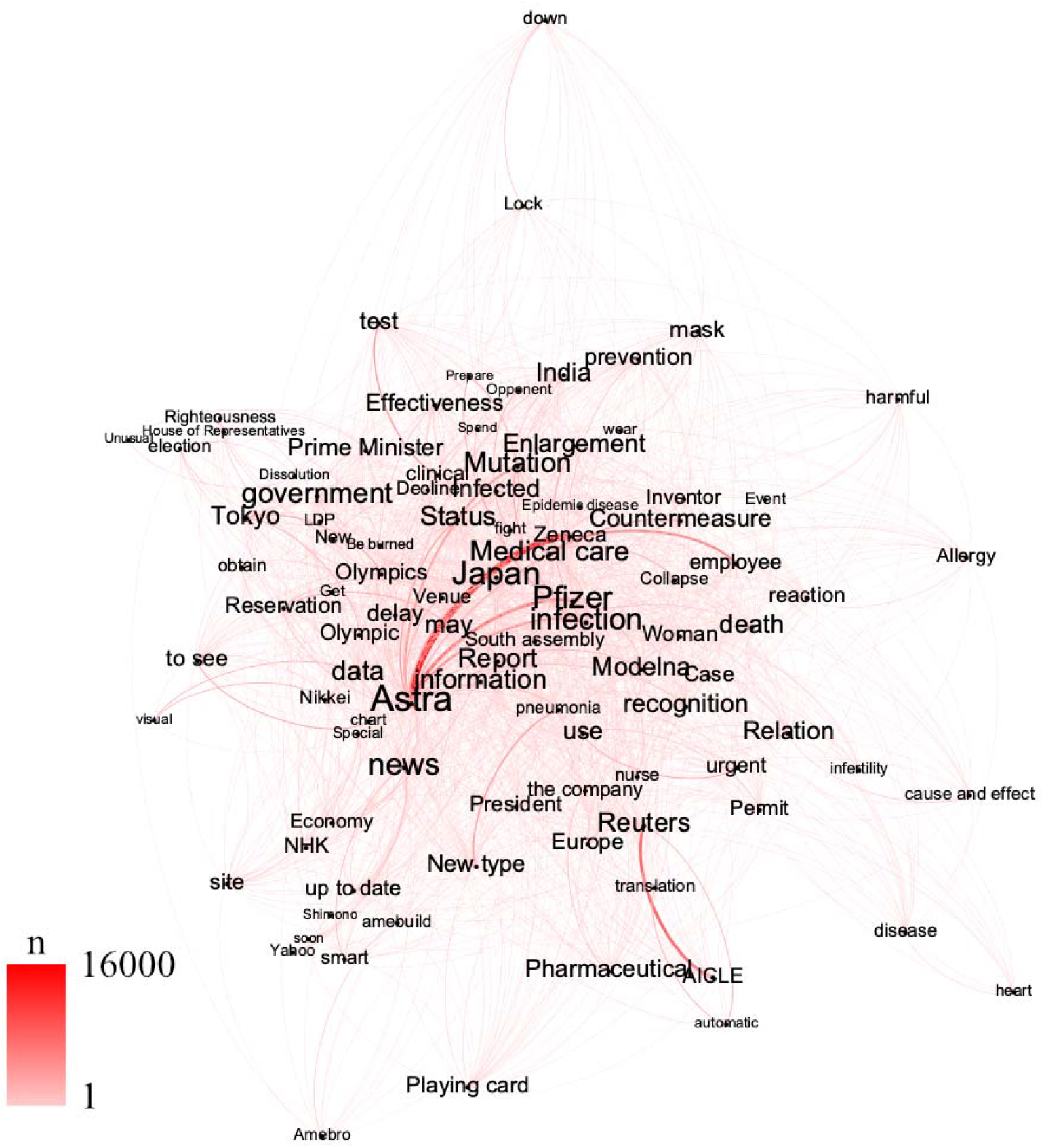
Network of top-100 bigram tokens in the corpus (English Version). The edges of the graph are the frequency of appearance of each token in the corpus, where thicker and more opaque edges indicate higher frequency.

### Topics and Sentiments towards the COVID-19 Vaccine

LDA modeling results are in Figure 4 (the word cloud is in Appendix 1 Figure 4), and a theme was summarized for each topic. Topic 1 is about vaccine appointment and distribution strategy; Topic 2 relates to the development progress of vaccines by different manufacturers and approval status of countries; Topic 3 concerns vaccine side effects and effectiveness against mutated viruses. We also provided the LDA modeling results of each sentiment in English as Appendix 1, Figure 5-12.

**Figure 4.**
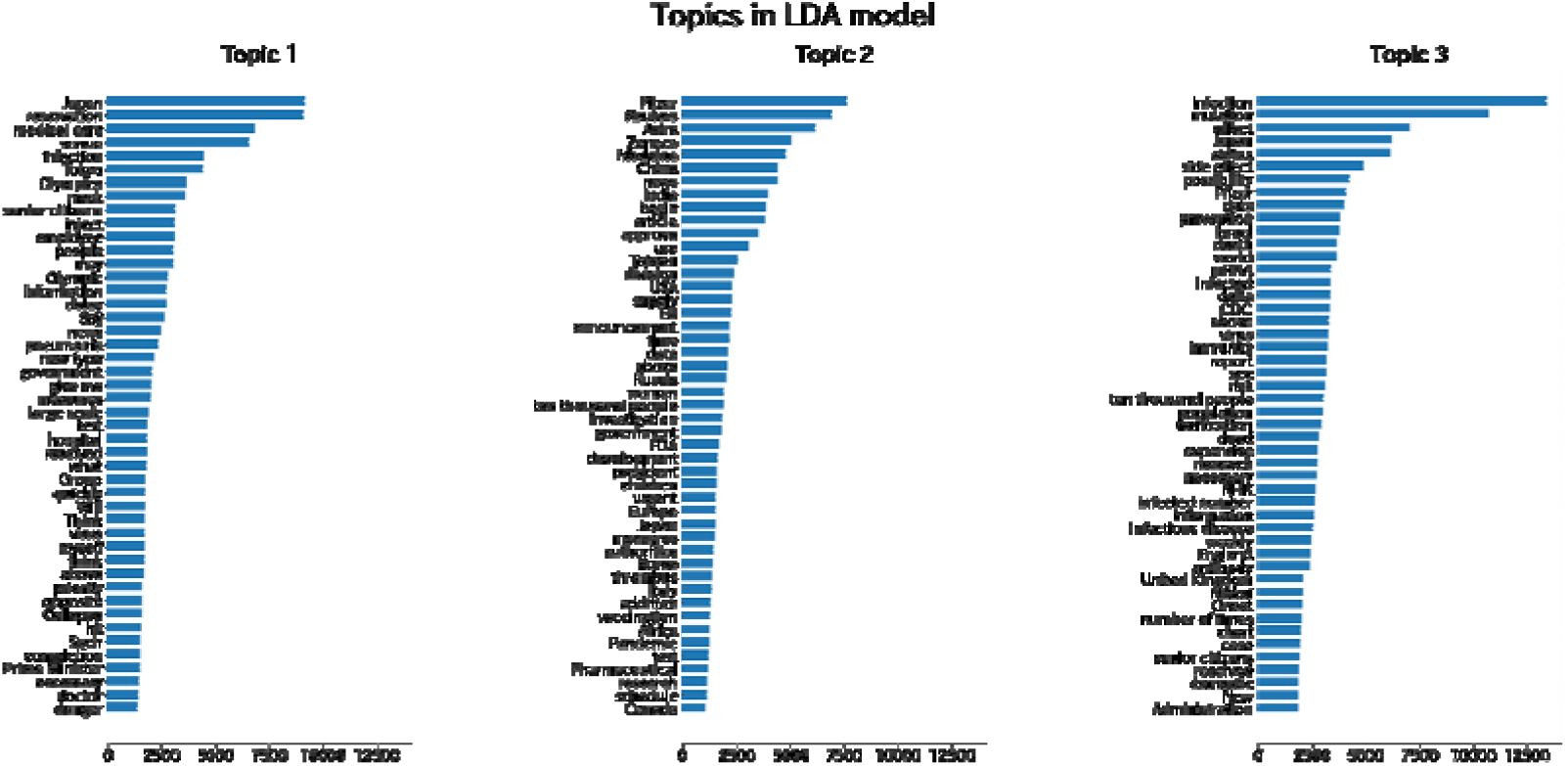
LDA topic modeling on all the vaccine-related tweets (English version).

We applied sentiment analysis to all tweets and counted the number of daily tweets for different sentiments in Figure 5. The sentiment was overwhelmingly neutral, with negative sentiment out volume positive sentiment. In Figure 6, we displayed the trends of positive and negative tweets with events. Negative sentiments mainly came from three aspects. The prompt response to vaccine rollout in other countries and news related to the side effects elicited negative public sentiment in Japan. Russia approved the first world’s COVID-19 vaccine (August 11, 2020, n=47); China started to give the self-developed vaccine for medical workers on August 25, 2020 (n=74). Negative news about vaccinations in other countries also caused negative sentiment in Japan. AstraZeneca and J&J paused on their studies on September 9, 2020 (n=49) and October 13, 2020 (n=42); A nurse in the US infected after the first dose vaccinations, on December 30, 2020 (n=127); WHO suggested that children should not be vaccinated in the current stage on June 22, 2021 (n=159). Negative sentiment also came from the severe infection situation in Japan and the local vaccination policy. Infection and serious illness cases both reached the highest record on January 21, 2021(n=88), ten prefectures extend the emergency statement on February 2, 2021(n=114); Senior citizens started to get vaccinations on April 13, 2021 (n=114). In contrast, the high efficiency of the vaccine and the start of large-scale vaccinations in Japan triggered positive sentiment. On November 9 (n=83) and November 16 (n=41), 2020, Pfizer and Moderna announced the vaccine’s effectiveness at over 90% <u>[39]</u> and 94.5% <u>[40]</u>, and on June 25, 2021 (n=42) vaccine effect against Delta variant was reported; On June 4, 2021 (n=61), the chief cabinet secretary announced the opening of vaccinations appointments for workplaces and universities starting from June 21. In addition, it is notable that the positive news about clinical trials also caused a peak of negative attitudes on November 9, 2020 (n=49).

**Figure 5.**
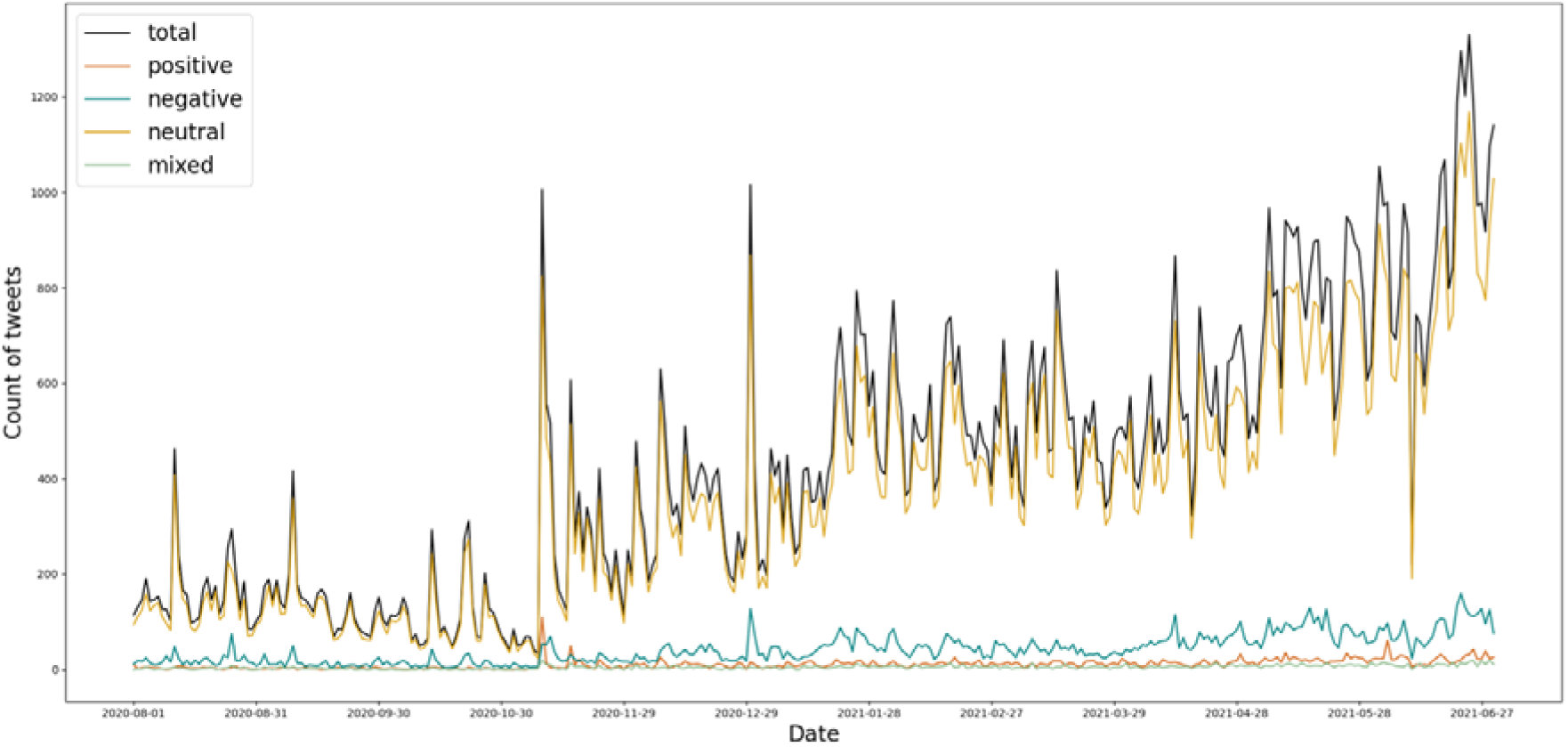
Trend of different sentiments between August 1, 2020, and June 30, 2021.

**Figure 6.**
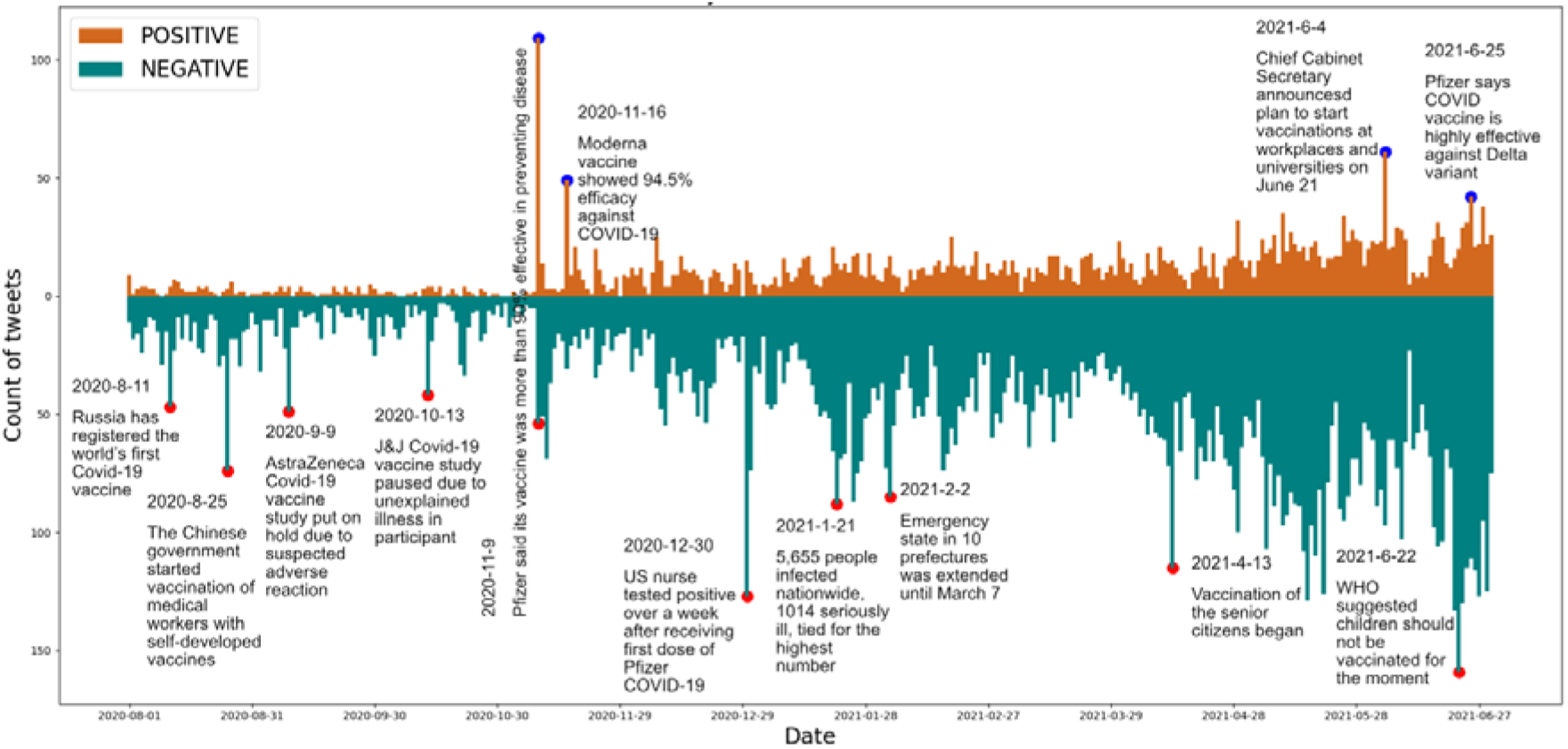
Trends of positive and negative sentiments between August 1, 2020, and June 30, 2021. The headline news related to peaks was labeled.

### Correlation of Total Tweets and Positive/Negative tweets with Infection, Death, and Vaccinated cases

As shown in Figure 7 and Figure 8, we calculated the correlation coefficients of total tweets and positive/negative tweets with the daily death, infection, and vaccinated cases respectively, both before and after the first vaccination in Japan (February 17, 2020, dashed line in Figure 8). As Table 2 shows, the daily number of tweets correlated with the number of deaths, infections, and vaccinations by 0.642, 0.405, and 0.686, respectively (0.715 after the first vaccination); Negative sentiment was strongly correlated with death (r=0.691) and infection (r=0.500) cases before the first vaccination, but they decreased to 0.305 and 0.293, respectively. The correlation between negative sentiment and vaccinated cases was slightly higher compared to positive sentiment after the first vaccination.

**Figure 7.**
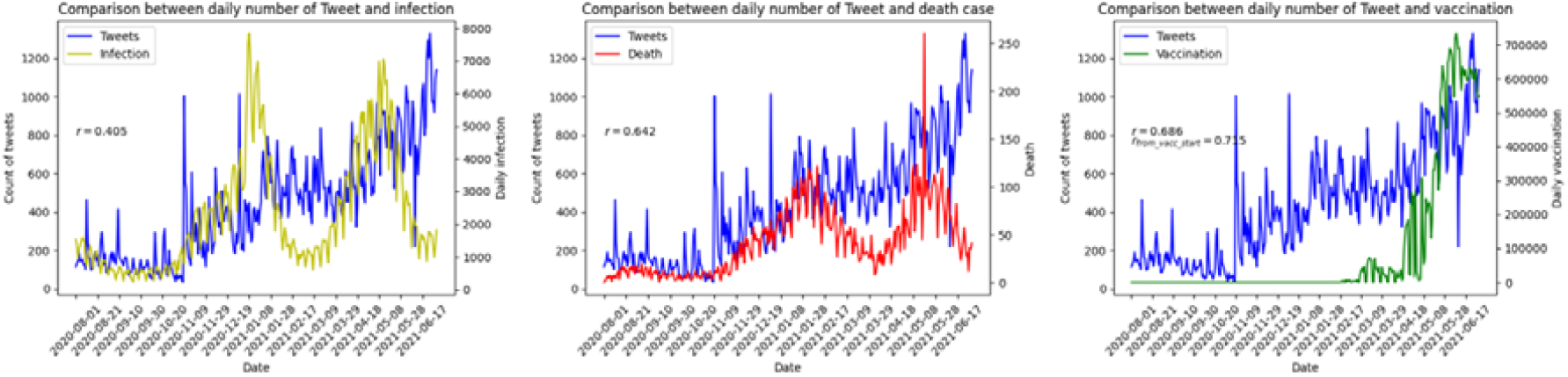
Trends of daily number of tweets with the daily number of death, infection, and vaccinated cases.

**Figure 8.**
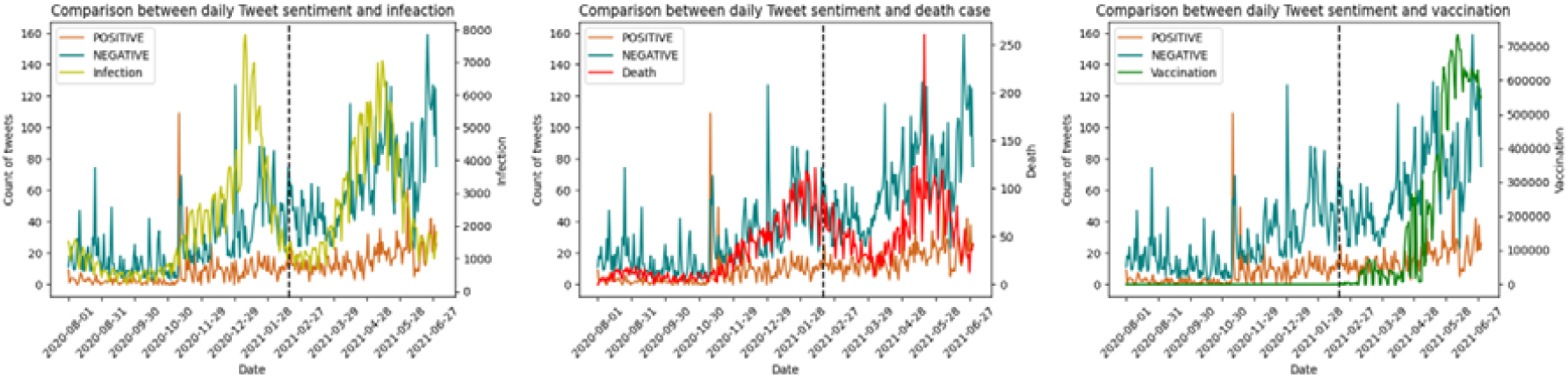
Trends of positive and negative sentiment together with the daily number of death, infection, and vaccinated cases. The dashed line indicates the first day of vaccinations in Japan (February 17, 2020).

**Table 2.**
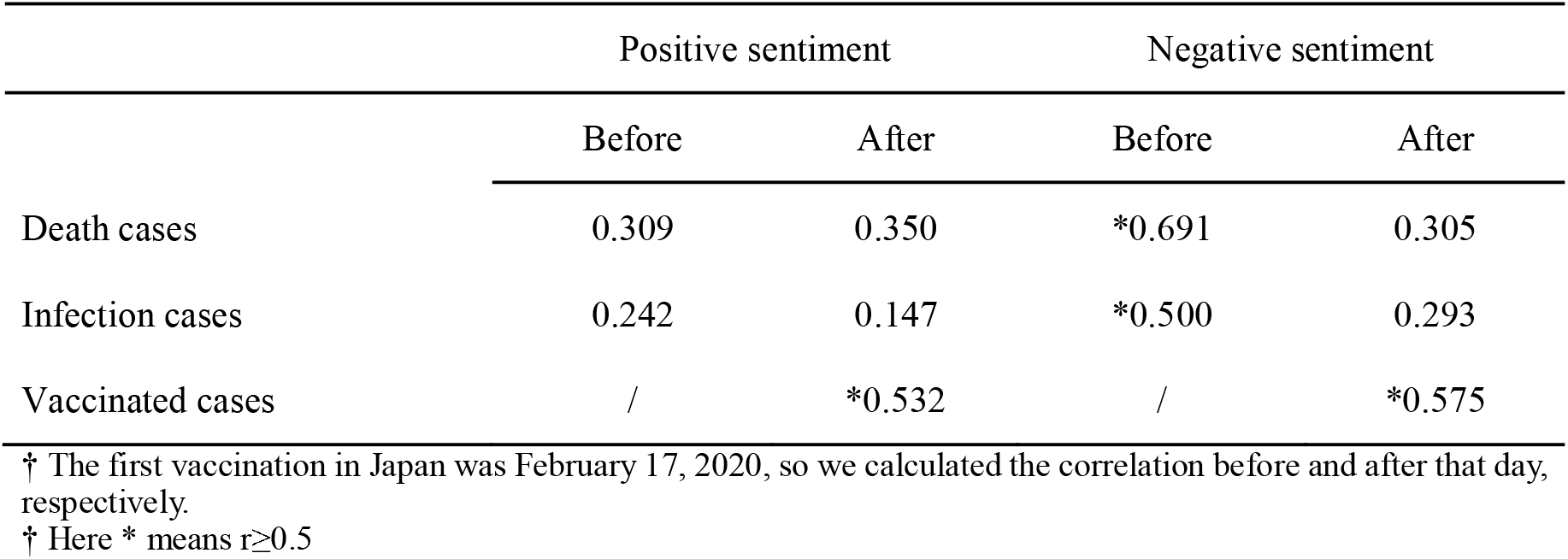
The correlation of the positive and negative sentiment with the daily death, infection, and vaccinated cases before and after first vaccination in Japan.

|  | Positive sentiment |  | Negative sentiment |  |
| --- | --- | --- | --- | --- |
|  | Before | After | Before | After |
| Death cases | 0.309 | 0.350 | *0.691 | 0.305 |
| Infection cases | 0.242 | 0.147 | *0.500 | 0.293 |
| Vaccinated cases | / | *0.532 | / | *0.575 |

† The first vaccination in Japan was February 17, 2020, so we calculated the correlation before and after that day, respectively.
† Here \* means $r \geq 0.5$

### Top Words of Three Manufactories Vaccines

Since June 30, 2021, the Japanese government has approved three vaccine brands. Tweets related to each vaccine brands were extracted, and the daily/monthly tendency of the three vaccine brands are shown in Figure 9. The numbers of related tweets, in descending order, are Pfizer (n=12,089), AstraZeneca (n=7,300), and Moderna (n=3,338), and the most representative news related to spikes in number of tweets was also consistent with Figures 1 and 6. Public concerns about the three vaccines had temporal variations. AstraZeneca vaccines showed an overall upward trend with peaks in September 2020 and March 2021, and a gradual decline after March. The Pfizer vaccine received greater attention than the others from October 2020 to February 2021 and reached a peak in November 2020. Moderna appeared less in discussions, but since March 2021, the number of related tweets has continued to grow along with the Pfizer vaccine.

**Figure 9.**
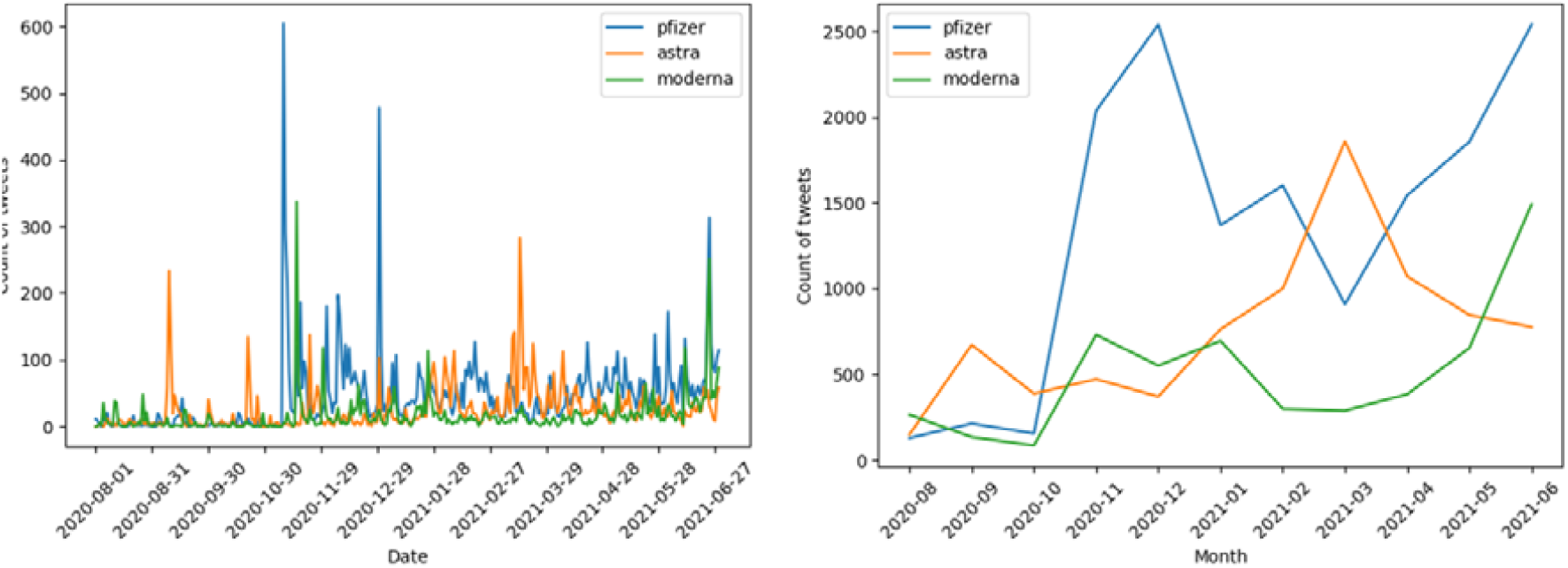
Tendency of the Pfizer, AstraZeneca. and Moderna vaccines between August 1, 2020, and June 30, 2021.

We analyzed the top 50 words in the tweets related to each vaccine brands. As shown in Figure 10 (the word clouds are in Appendix 1 Figure 13), the discussion of all three vaccines was surrounding the keywords “Japan,” “approve,” “effect,” “mutation,” “clinical trials,” “supply,” and “side effects”. Pfizer, the first vaccine approved by the Japanese government, was discussed more than the sum of AstraZeneca and Moderna. We also checked the LDA for the three vaccine brands, as shown in Appendix 1 Figure 14-19. However, the daily number of tweets was too small, and it was difficult to perform further analysis according to the results.

**Figure 10.**
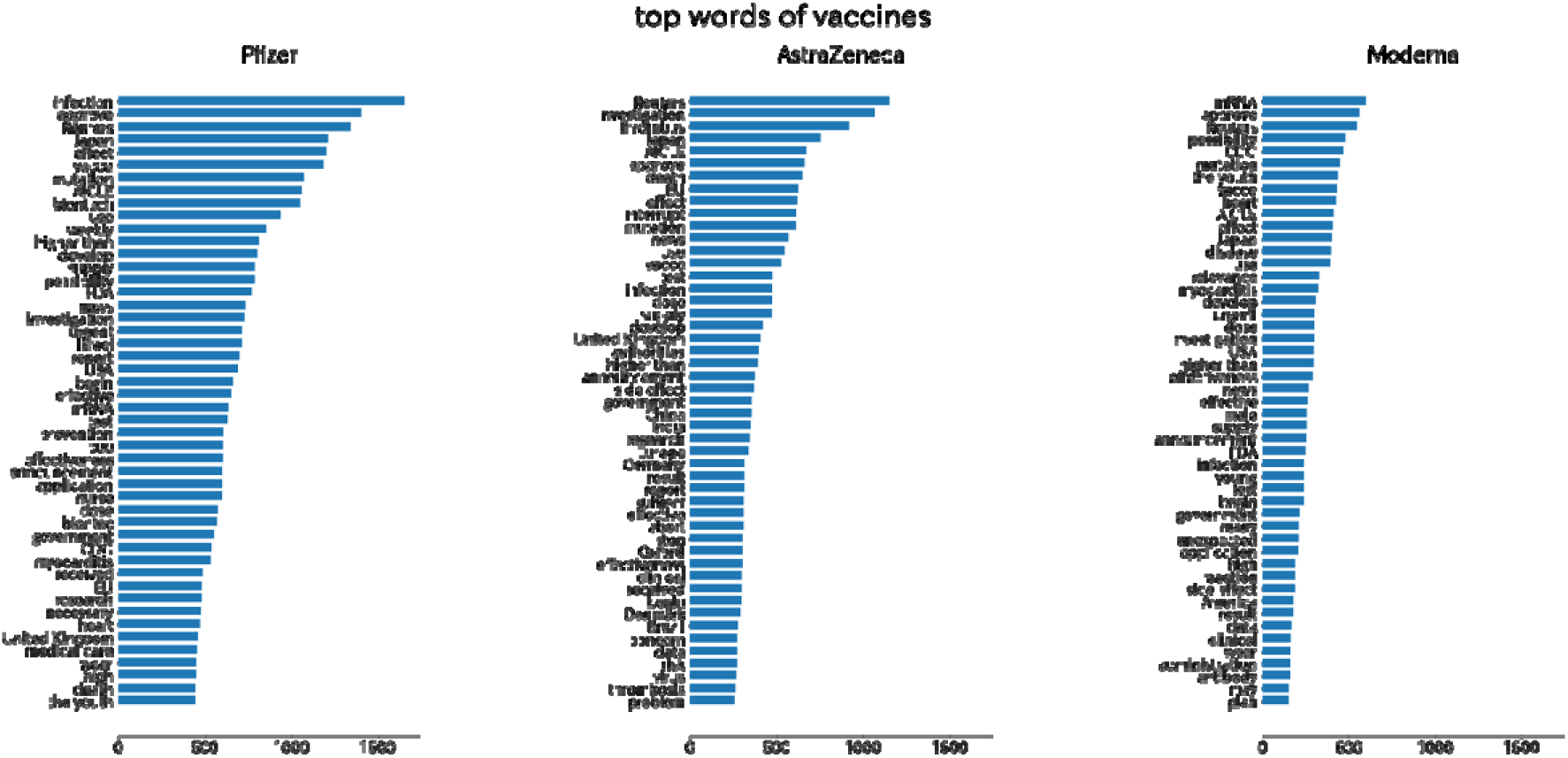
Top words associated with the Pfizer, AstraZeneca, and Moderna vaccines. The length of the bar indicates the number of times a word occurs.

## Discussion

### Principal Findings

This study examined long-term Japanese public opinion and sentiment, covering discussions from August 1, 2020, through June 30, 2021, when multiple vaccines were available, yet only 13.6% of the population was fully vaccinated in Japan. We evaluated the tendency of number of Japanese tweets and identified the unigram and bigram keywords. Overall number of tweets continued to increase since the start of large-scale vaccinations in Japan, which might be primarily driven by critical events related to vaccines. We also checked the LDA topics and temporal changes of sentiment. The public concerns include three topics: vaccine appointment and distribution strategy; Development progress of different manufacturers and approval status of countries; Vaccine side effects and effectiveness against mutated viruses. About 85% tweets were of neutral sentiment, and negative sentiment overwhelmed positive sentiment in the other tweets. Additionally, we calculated the correlation between the number of tweets/positive/negative sentiment and infection/death/vaccinated cases. The results revealed infection/death cases correlated strongly with negative sentiments, but the correlation weakened after vaccinations rollout as infection/death cases decreased. Eventually, we analyzed public sentiment about three vaccine brands (Pfizer, Moderna, and AstraZeneca), which had a temporal shift as clinical trials moved forward, but the core remained effective and secure.

Our results showed that negative sentiment predominates positive sentiment in most time in Japan, when most previous studies of other countries exhibited more positive sentiment on social media [23,24,26]. The negative sentiment exhibited by our results consistent with the findings of some previous studies using surveys in Japan [40–45], and we provided fine-grained and more practical evidence. We observed a decrease in the correlation of negative sentiment with infection cases and death cases in Table 2, for which the direct reason is the decrease of infection and death cases and the abnormal increase of negative sentiment. The decrease of infection and death cases may result from the effect of the emergency statement and vaccinations, but the increase of negative sentiment may come from the frequent negative news about the messy process of vaccinations including busy phone lines, websites crashing, and incorrectly administered vaccinations in this period. We also found the same event could trigger positive also negative sentiments in Japan. On November 9, 2020, when Pfizer reported its vaccine was more than 90% effective, the peak of both positive and negative sentiments may indicate the public expectation of its effectiveness and concerns about its safety. We presume the negative sentiment was caused by the accumulation of negative news from different vaccines during clinical trials. downward trend in both deaths and infections after the first dose, contrary to the trend of a consistent increase in the number of tweets.

The LDA topics in our study reflected the public concerns towards vaccinations and the corresponding policies. Topic one of LDA is about vaccine appointment and distribution strategies. Although vaccine appointments opened in Japan, the government decided to give priority appointments to healthcare workers and seniors over 65 years old. However, this policy generated widespread negative sentiment, with some Twitter users arguing that priority should be given to those in the public service sector rather than to senior citizens, who have less contact with people. In contrast, Indonesia prioritizes vaccinations for those of productive age (18-59 years) over the seniors [46]. Matsui and colleagues also suggested equal distribution from public policy and economic perspectives [47]. Sunohara and colleagues suggested that the current strategy is better only if a smaller economic loss and a stronger lockdown [48]. Topic two indicated that the public focused on vaccine development by different manufacturers and approval status by country. Due to slow implementation, Japan ranked the worst among the organization for Economic Cooperation and Development member countries. Also, probably because Japan completely relied on vaccine imports, the public paid attention to the approval and vaccination status of other countries. Topic three showed that the public was concerned about side effects and effectiveness against mutated viruses. The slow implementation of vaccinations in Japan became major trouble against the epidemic. Coupled with mutated viruses spread across multiple districts, the Japanese public particularly cared about the effectiveness of vaccines against mutated viruses.

The opinions and sentiments towards different vaccine brands are slightly different, but the core remained effective and secure. Our analysis of the three vaccines (Pfizer, Moderna, and AstraZeneca) shows that public opinions and concerns about different types of vaccines are consistent in Japan, such as effectiveness, availability, vaccination information, and side effects. Compared to the other two vaccines, the public tended to focus on the effectiveness of Pfizer in preventing infection, as opposed to Moderna, which tended to focus more on its effectiveness against mutated viruses and its mRNA development technology. Furthermore, we found that “fake news” or misleading headlines caused public panic and widespread negative sentiment. For example, on October 21, 2020, numerous media reported that the clinical trial of AstraZeneca vaccine led to the death of a Brazilian volunteer, which continued to trigger public panic even though the next day it was reported that the volunteer did not receive the vaccine[49]. In an information-exploding network, we regard mainstream media as being responsible for ensuring objective and accurate news and recommend that authorities should actively and quickly respond to inaccurate reports and fake news to establish public confidence in vaccines and build scientific understanding.

### Implications and Recommendations

The popularity of social media platforms coupled with natural language processing strategies benefits the government by enabling the monitoring of close-to-real-time public sentiment towards vaccine information. This can inform more effective policymaking and establish confidence towards vaccines, to maximize vaccine uptake. Some of our findings provide new evidence and inspiration to academic researchers and can assist policymakers to capture the relevant information needed in real-time.

For academic researchers, this study is the first application of sentiment analysis via social media related to healthcare in Japan to our knowledge, thereby supporting a feasible example for Japanese sentiment analysis. As of October 2021, Japan ranked second in the world with more than 58 million users[50], providing an enabling environment for opinion mining. However, its application remains nascent in Japan, especially in public health, although social media has been widely used by academia as a powerful tool. Moreover, we are deeply aware of the limited Japanese resources in text processing, our study may draw the attention of academia to develop more usable Japanese models for further research.

For policymakers, our study provides Japanese public opinion and sentiment towards the COVID-19 vaccine with dynamic and unmodified expression. Japan ranked among the countries with the lowest vaccine confidence in the world [10], which might be linked to the HPV vaccine safety scares that started in 2013. However, the way in which the HPV vaccine scare was approached by health officials, indicates continuing issues with the Japanese vaccination program that need resolving. Correspondingly, our findings indicate that the Japanese public showed significant negative sentiment before and at the beginning of vaccinations, in contrast to other countries [23,24,26]. To this end, we explored the Japanese public’s concerns about vaccines through LDA modeling and word frequency analysis and investigated the Japanese public’s perceptions of three different brands of vaccines, so that policymakers can fully understand public opinions and tailor more reasonable administration strategies.

### Limitations

Our study has several limitations. Firstly, Twitter penetration is only 58.2 million (42.3% of the total population) in Japan [27], our data may not be representative of the entire population, especially the senior generations [51]. Secondly, according to Twitter’s user privacy protection principles, our study did not further examine the demographic characteristics of users, such as age, gender, and geographic location. Thirdly, due to the limitations of Japanese resources for sentiment analysis, we used AWS without any training on the data, our findings may be influenced by the accuracy of the model. Eventually, only a small number of people got vaccinated during our study period, including senior citizens and healthcare workers. So, most of the tweets posted by users were based on the information from the internet and news, rather than direct experience with vaccinations. Accordingly, further in-depth studies are needed in the future.

## Conclusions

This study identified the Japanese public opinions and sentiments expressed on Twitter vaccine before and at the beginning of large-scale vaccinations, and their changes over time, with the aim of potentially facilitating vaccinations. It was suggested that these changes might be driven by critical news events related to the COVID-19 vaccine. The same event can induce opposite sentiment peaks, and inaccurate news may trigger public panic and distrust of vaccines. Japanese public presented more negative sentiments than positive ones, with concerns about the vaccine’s appointment availability, distribution strategy, vaccinated status of other countries, effectiveness, and side effects. We also found that the public attention to different vaccine brands remained effectiveness and safety, despite the discussion differing slightly. The policymakers should provide more evidence about the effectiveness and safety of vaccines and optimize the process of large-scale vaccinations.

## Data Availability

The authors provided all the raw datasets as supporting information to replicate our study findings.

http://www.panacealab.org/covid19/

## Ethical Approval

No ethical approval was required.

### Abbreviations

LDA: Latent Dirichlet Allocation
AWS: Amazon Web Services
HPV: human papillomavirus
NLP: natural language processing

