## Appendix1_Figures for "Public Opinion and Sentiment Before and at the Beginning of COVID-19 Vaccinations in Japan: Twitter Analysis"

Figure 1 Plot of number of topics against LDA tuning scores.

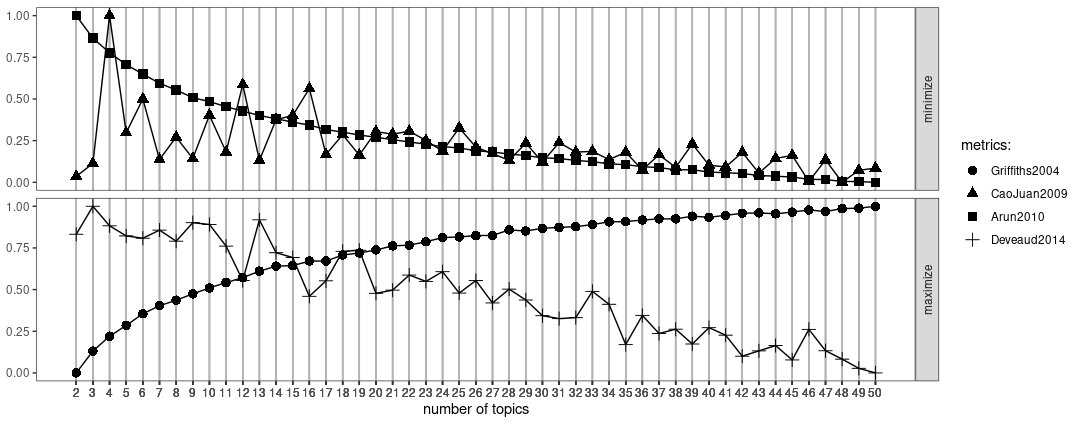

Figure 2 Count of monogram (unigram) tokens. The horizontal axis is the index of tokens by the frequency of appearance and the vertical axis is the count of appearance of the tokens.

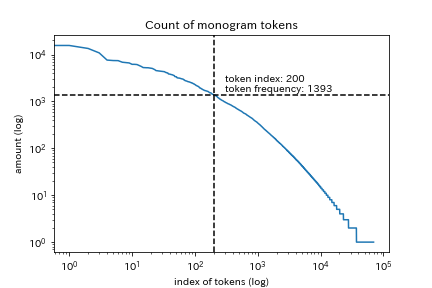

Figure 3 Count of bigram tokens. The horizontal axis is the index of tokens by the frequency of appearance and the vertical axis is the count of appearance of the tokens.

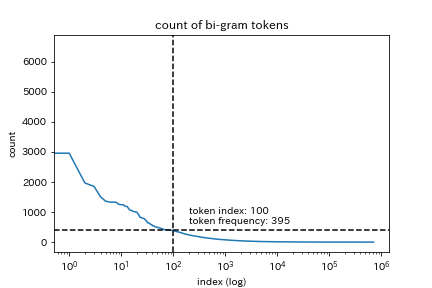

Figure 4. Word cloud of three LDA topics for all vaccine-related tweets (English version).

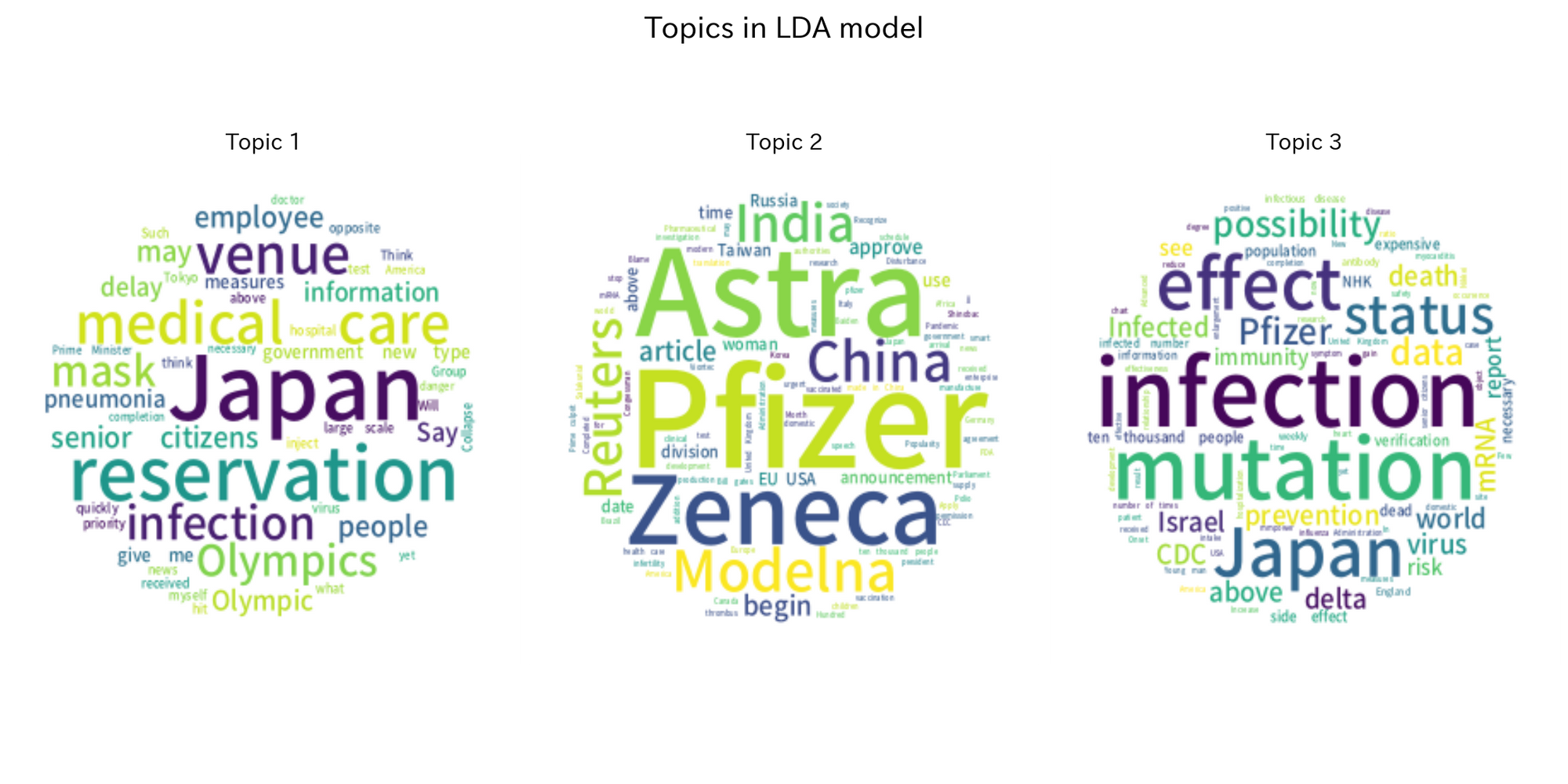

Figure 5. Top-50 words of three LDA topics for positive tweets (English version).

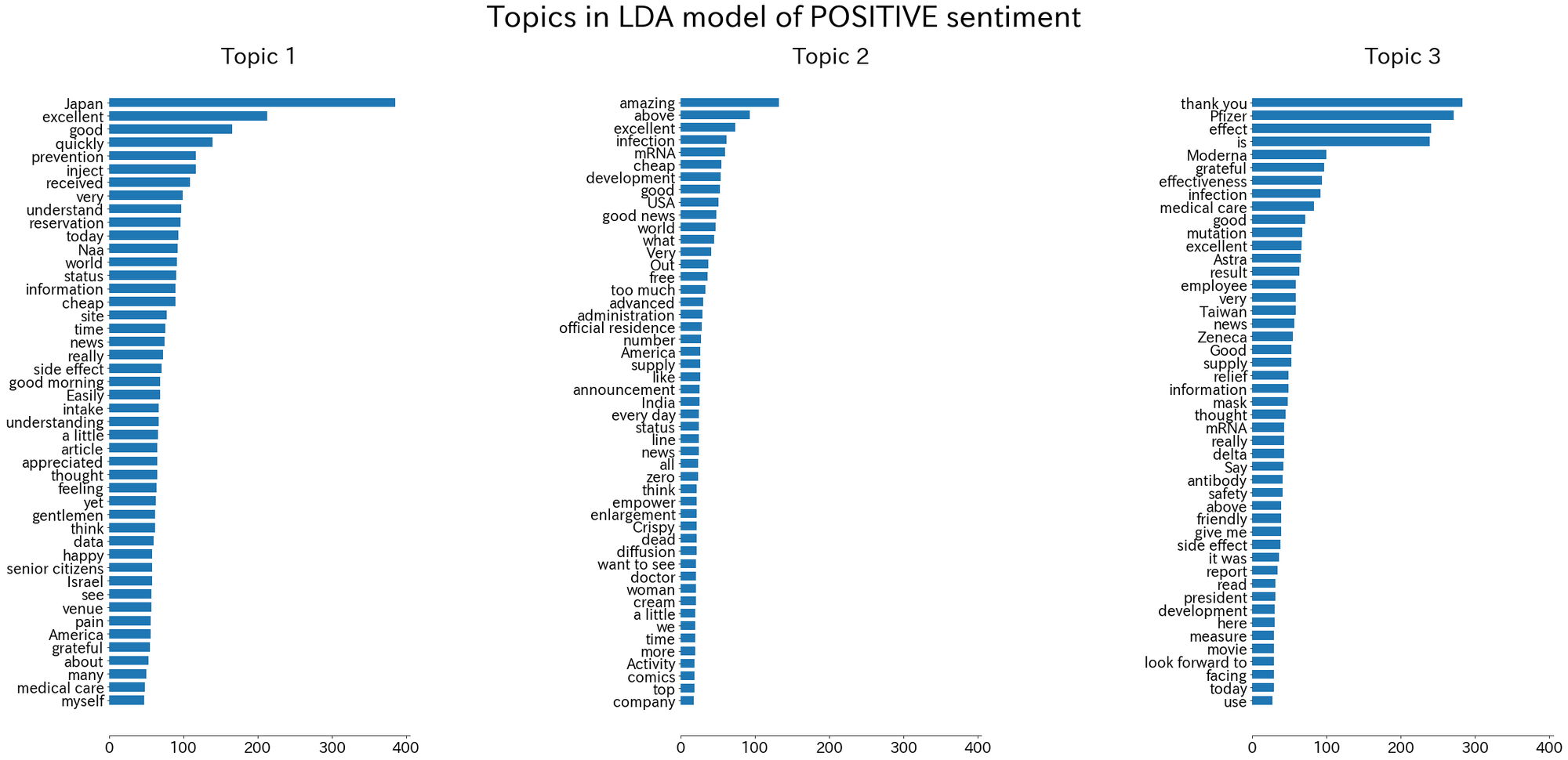

Figure 6. Word cloud of three LDA topics for positive tweets (English version).

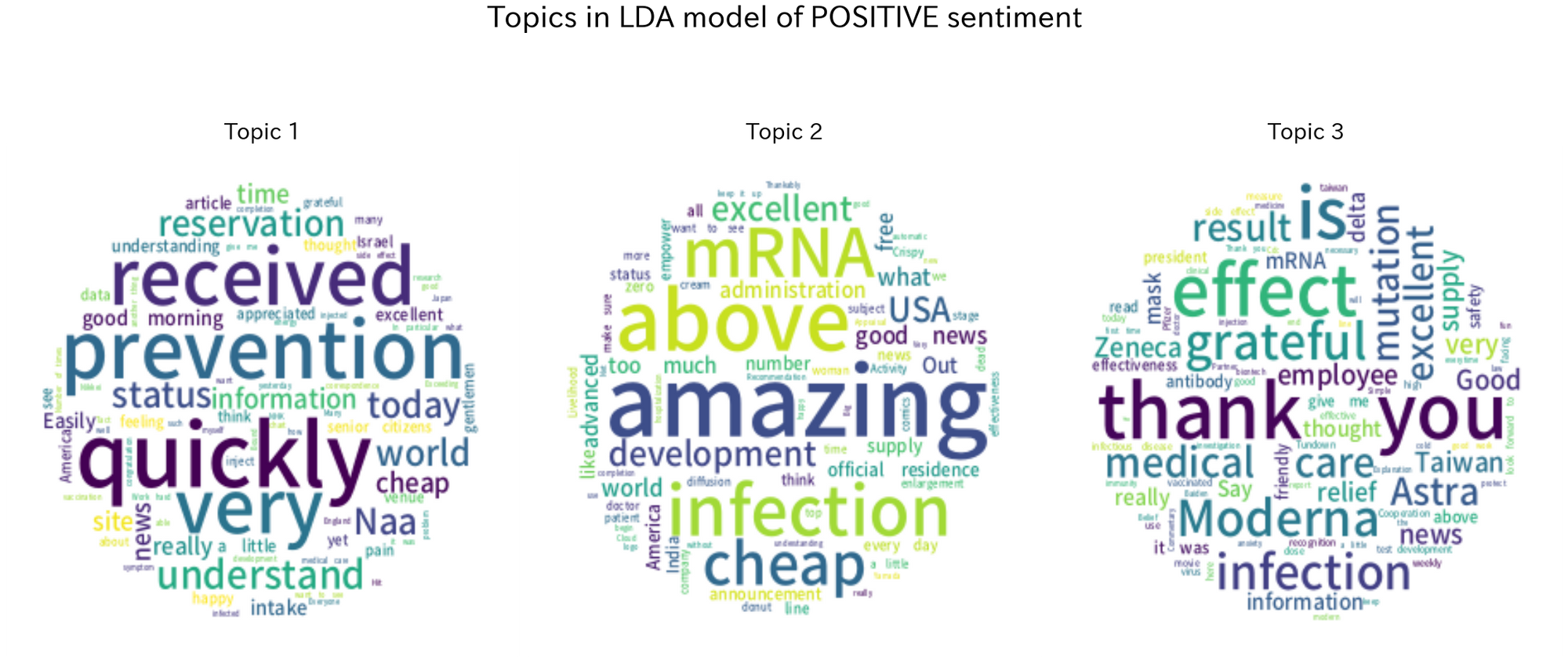

Figure 7. Top-50 words of three LDA topics for negative tweets (English version).

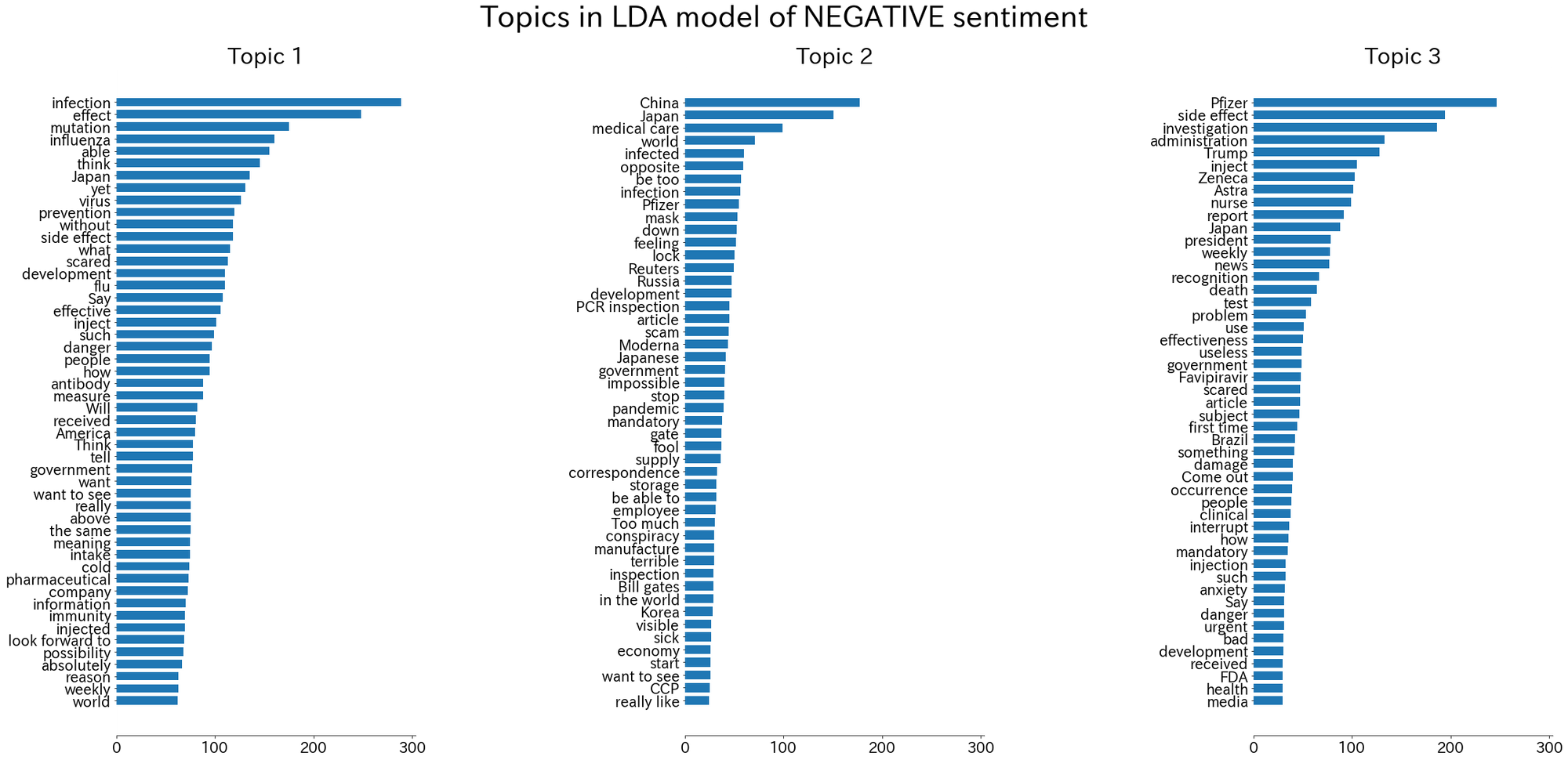

Figure 8. Word cloud of three LDA topics for negative tweets (English version).

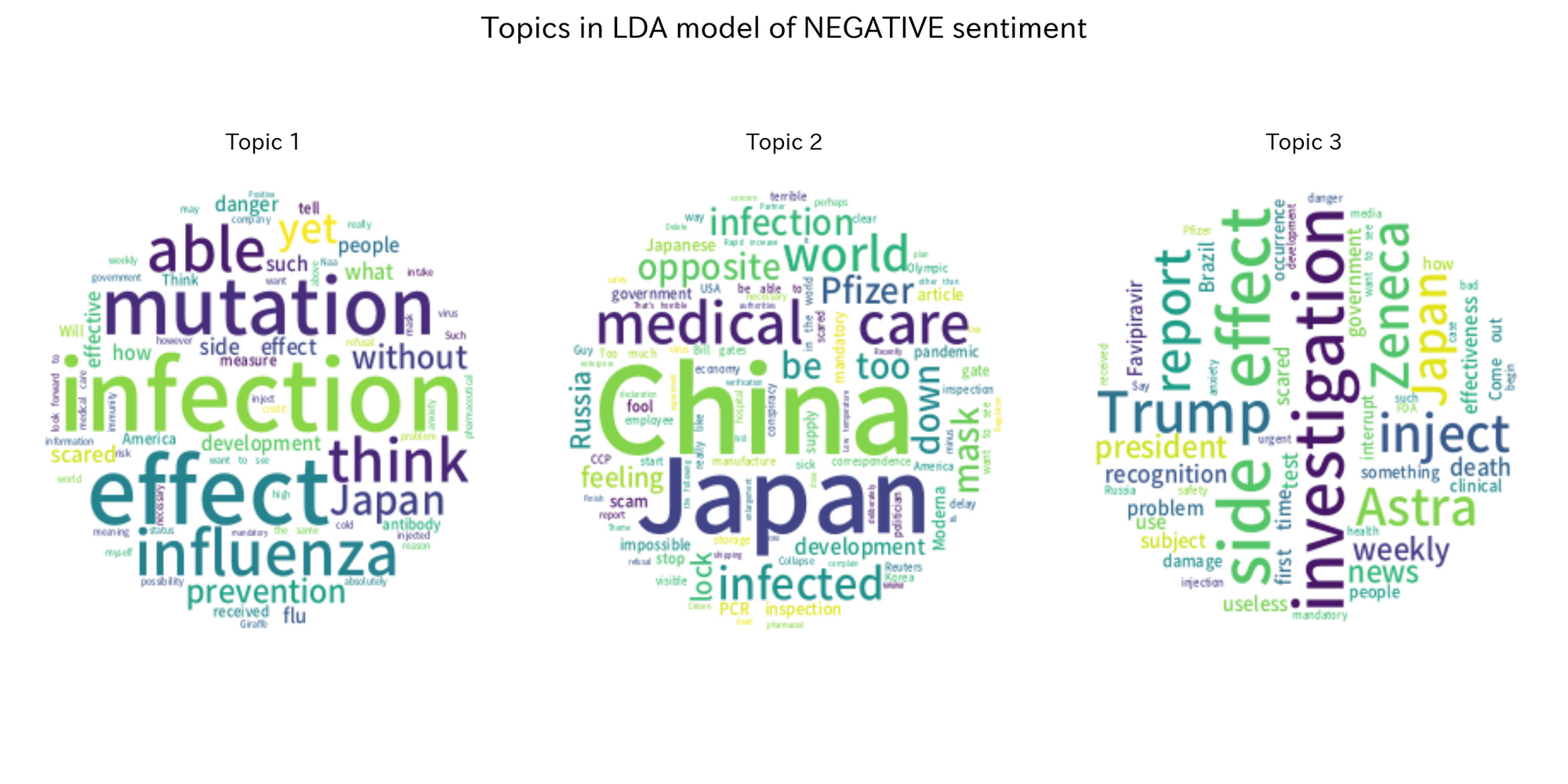

Figure 9. Top-50 words of three LDA topics for neutral tweets (English version).

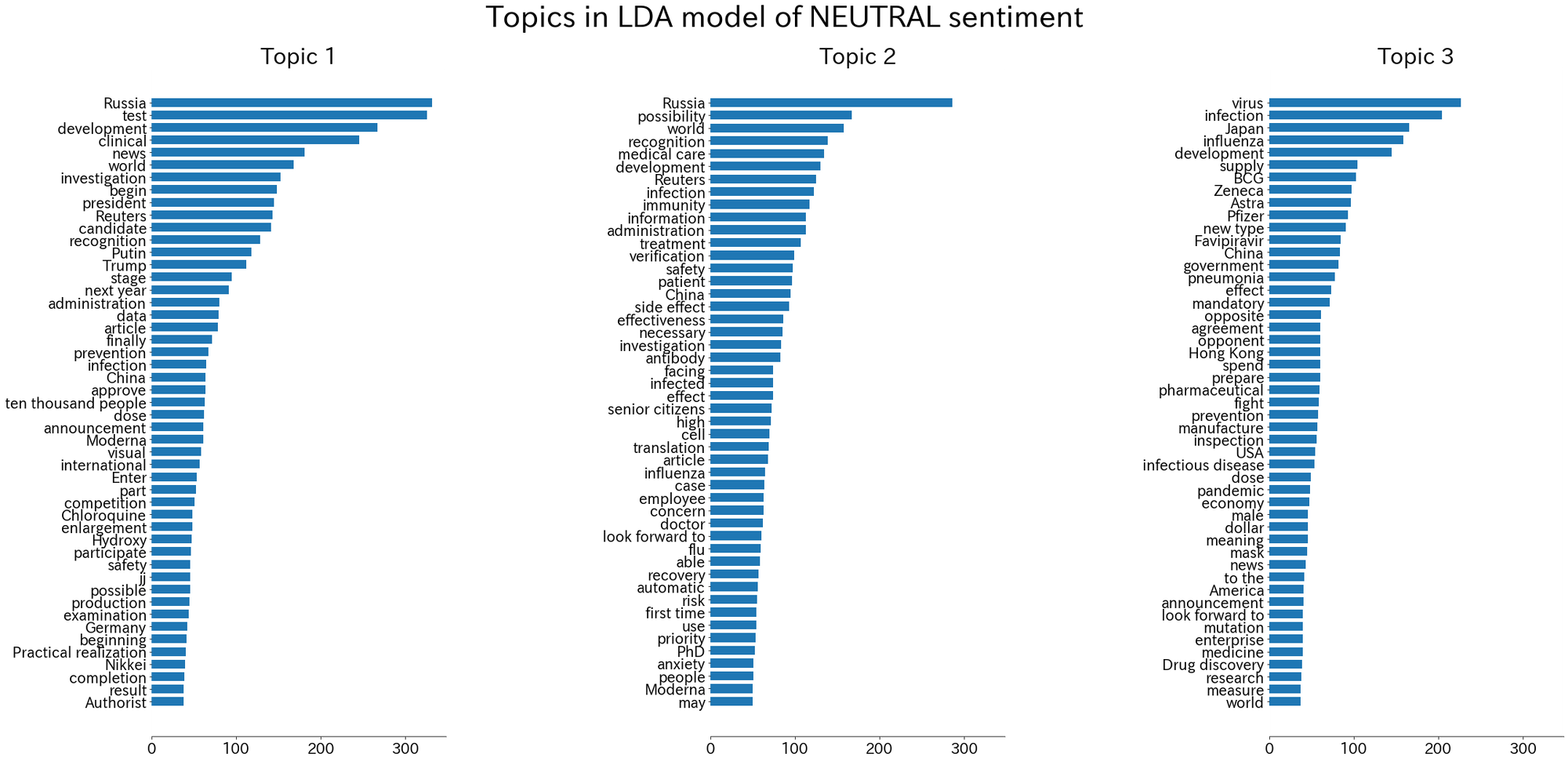

Figure 10. Word cloud of three LDA topics for neutral tweets (English version).

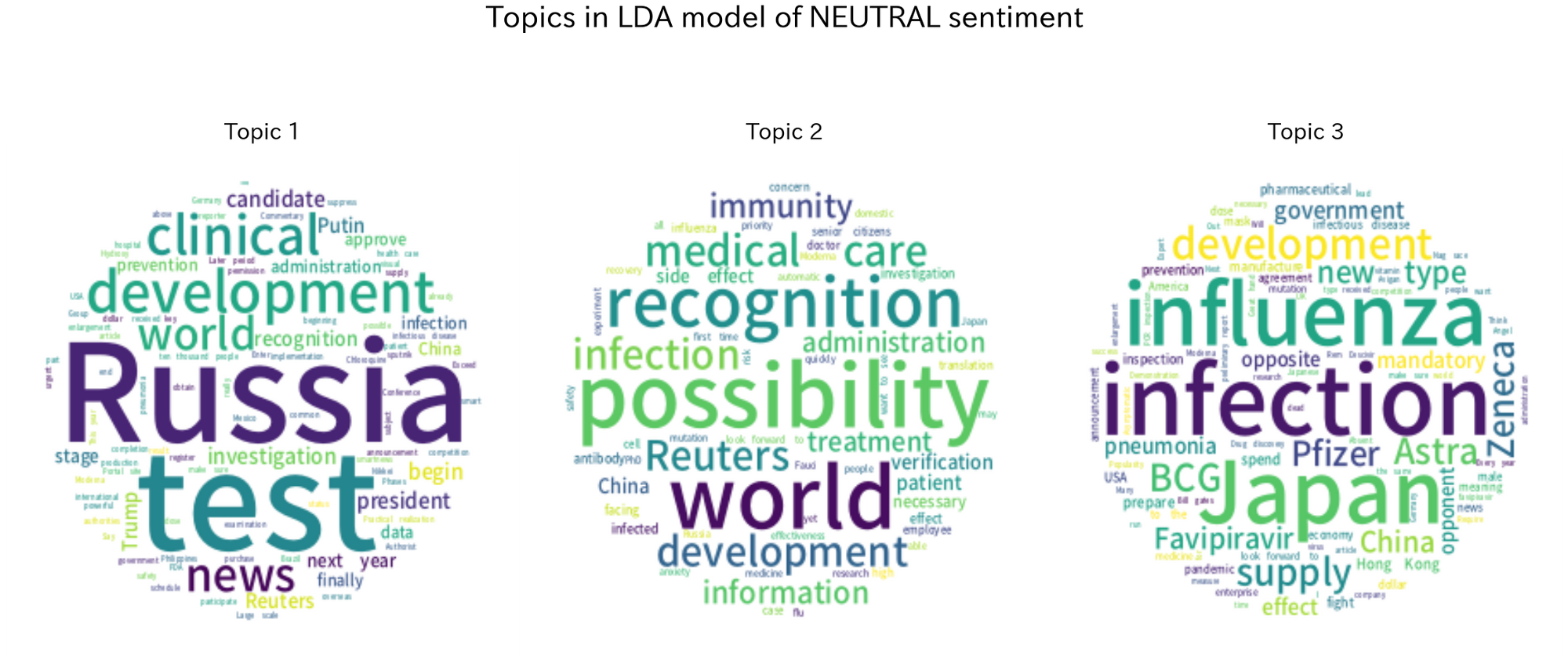

Figure 12. Word cloud of three LDA topics for mixed tweets (English version).

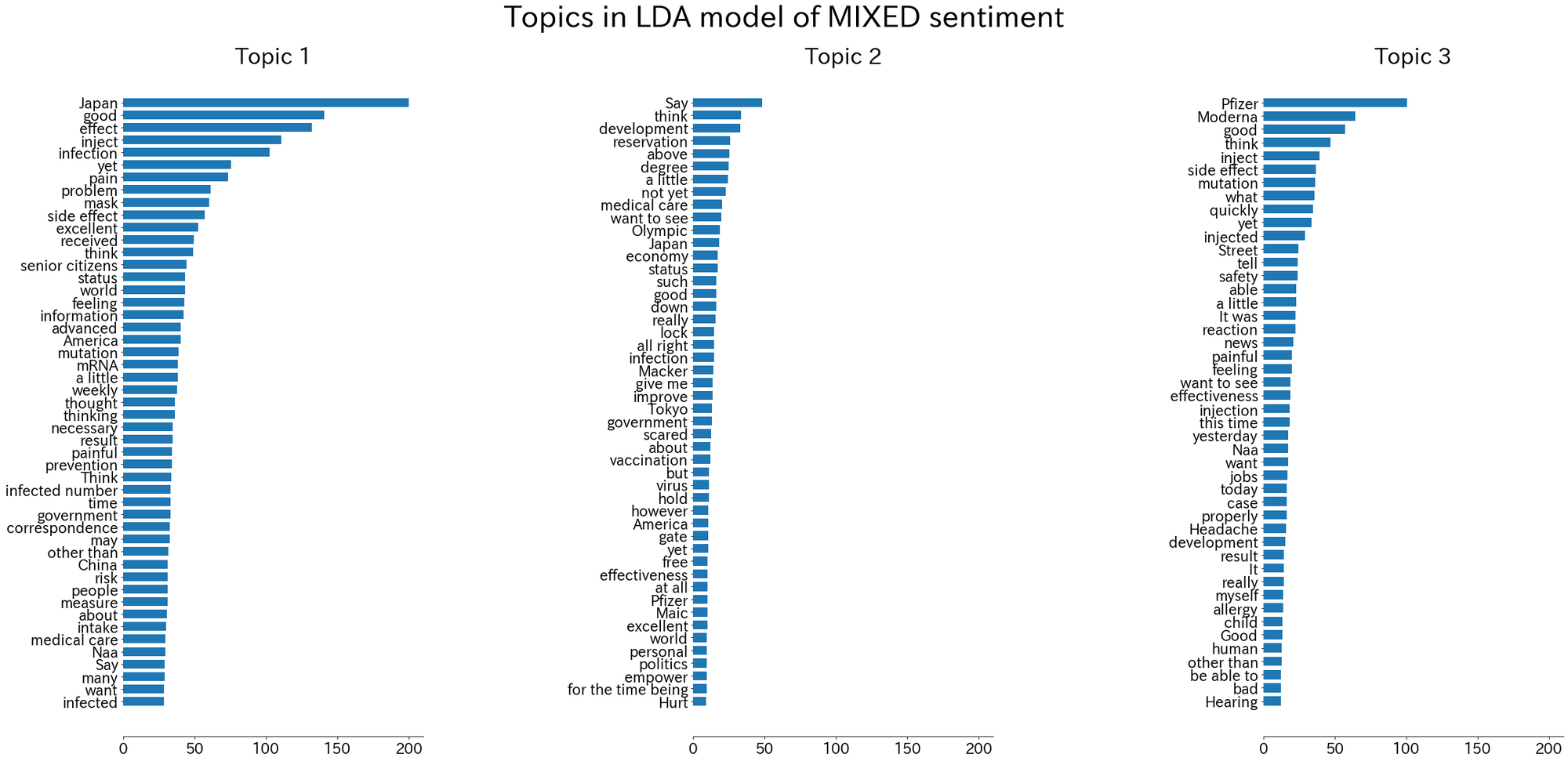

Figure 12. Word cloud of three LDA topics for mixed tweets (English version).

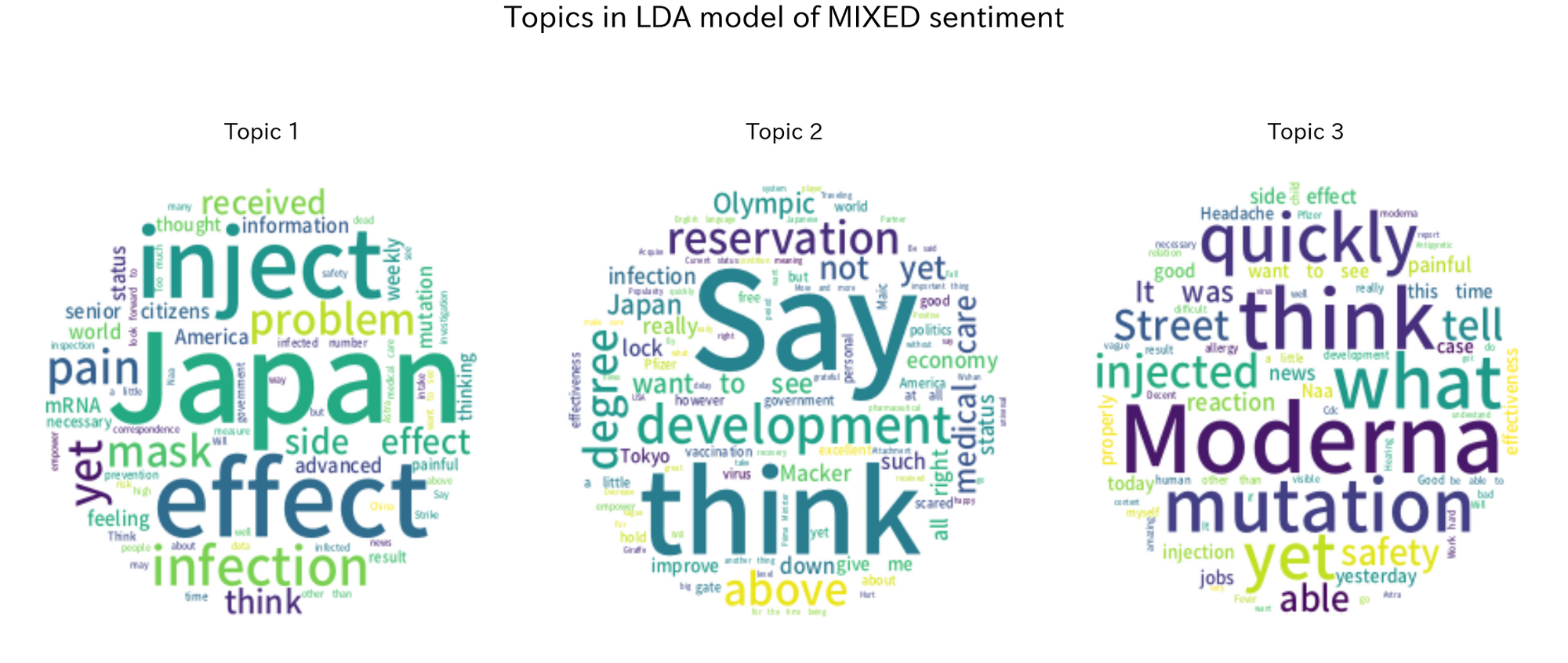

Figure 13. The top-50 words for each vaccine brand.
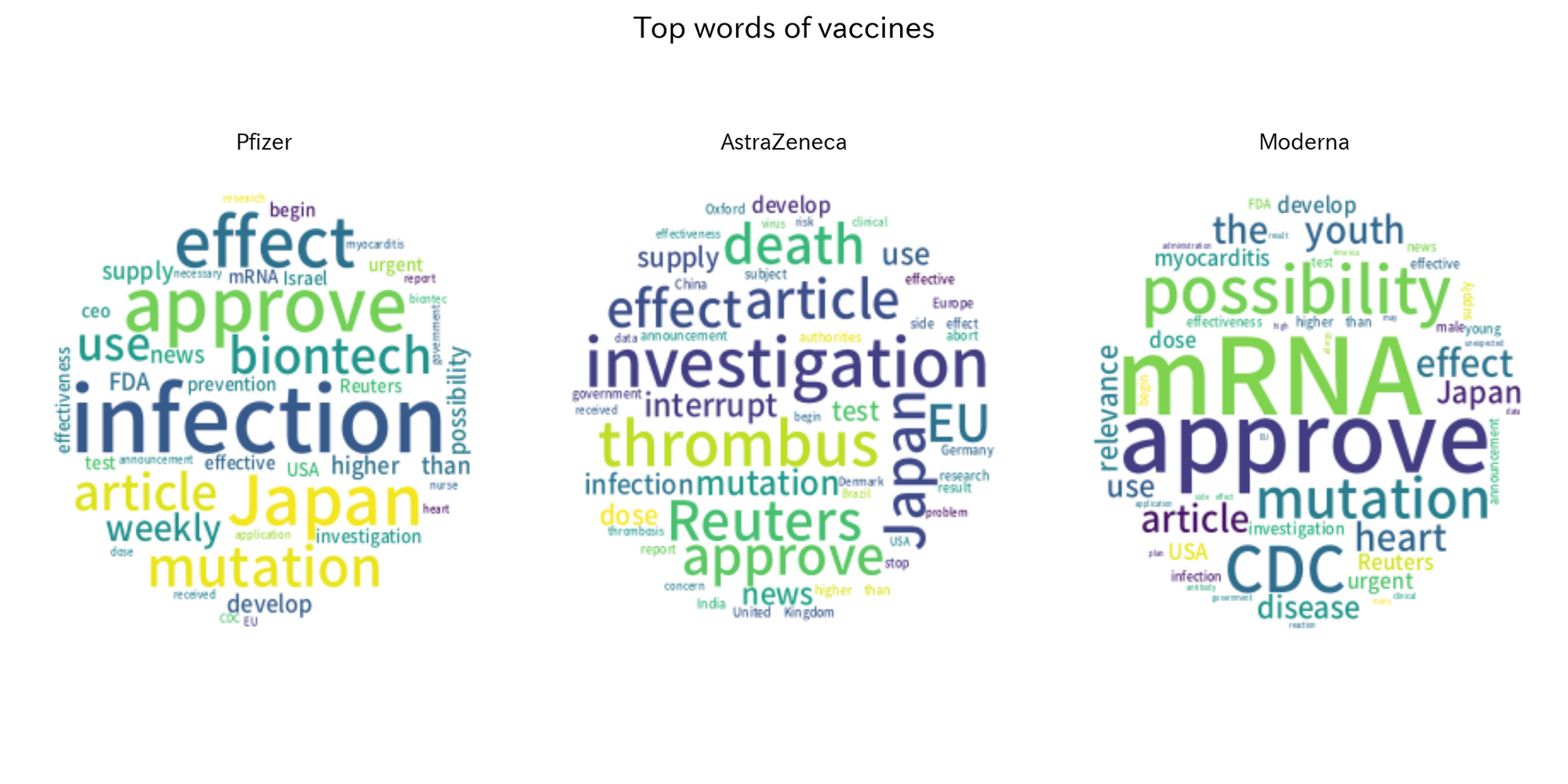

Figure 14. Top-50 words of three LDA topics for tweets related to Phizer vaccine (English version).

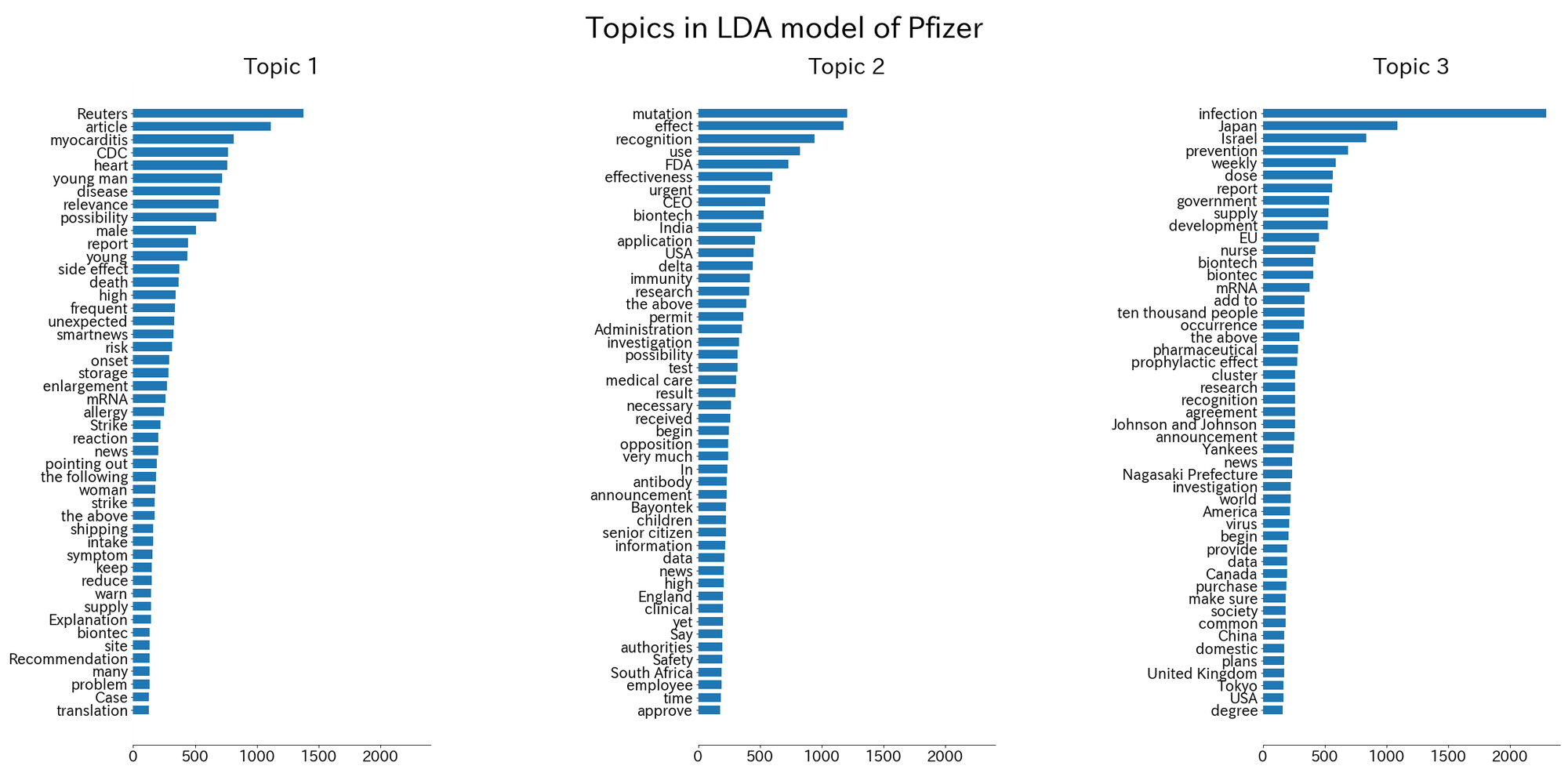

Figure 15. Word cloud of three LDA topics for tweets related to Phizer vaccine (English version).
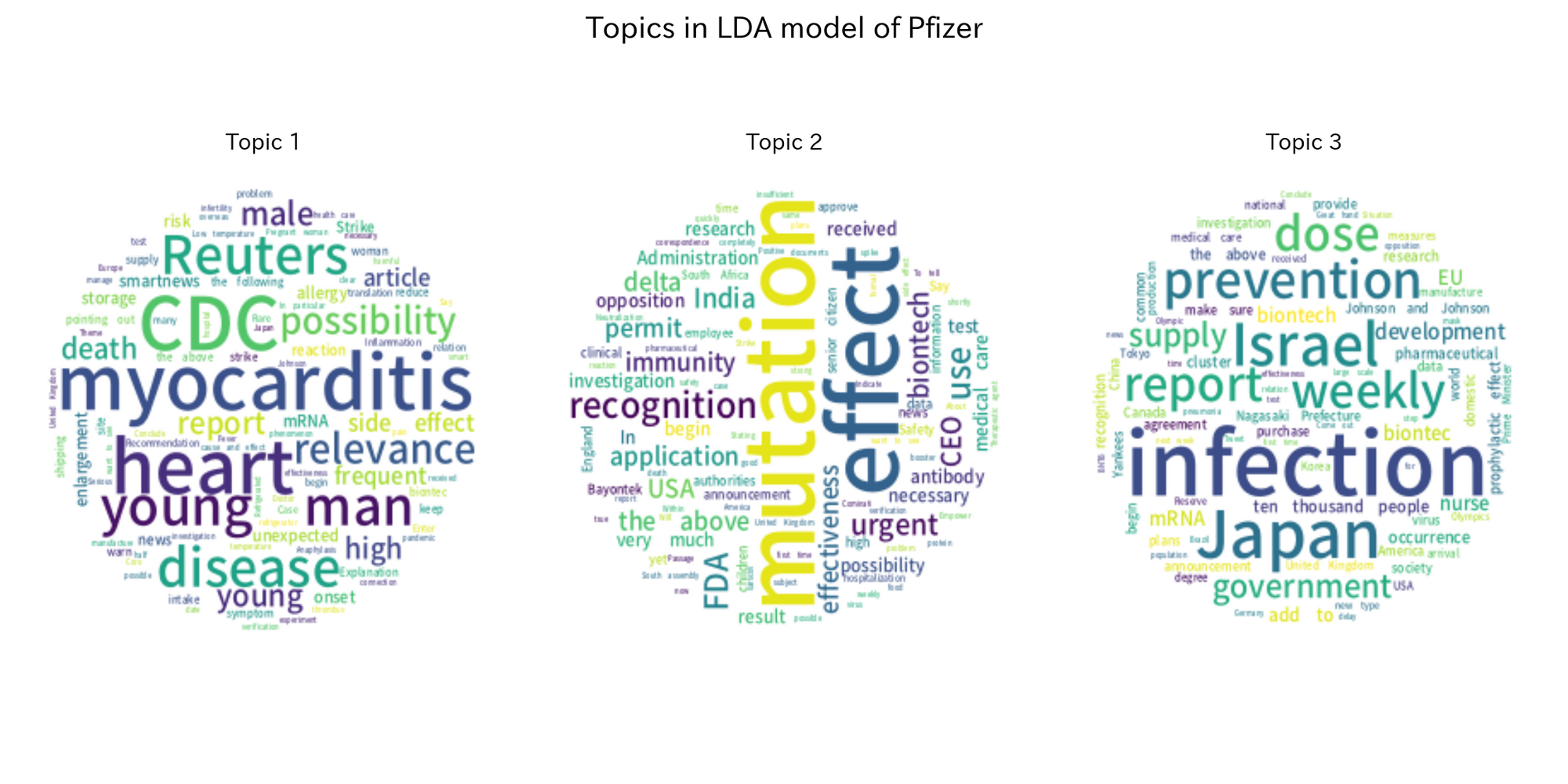

Figure 16. Top-50 words of three LDA topics for tweets related to AstraZeneca vaccine (English version).

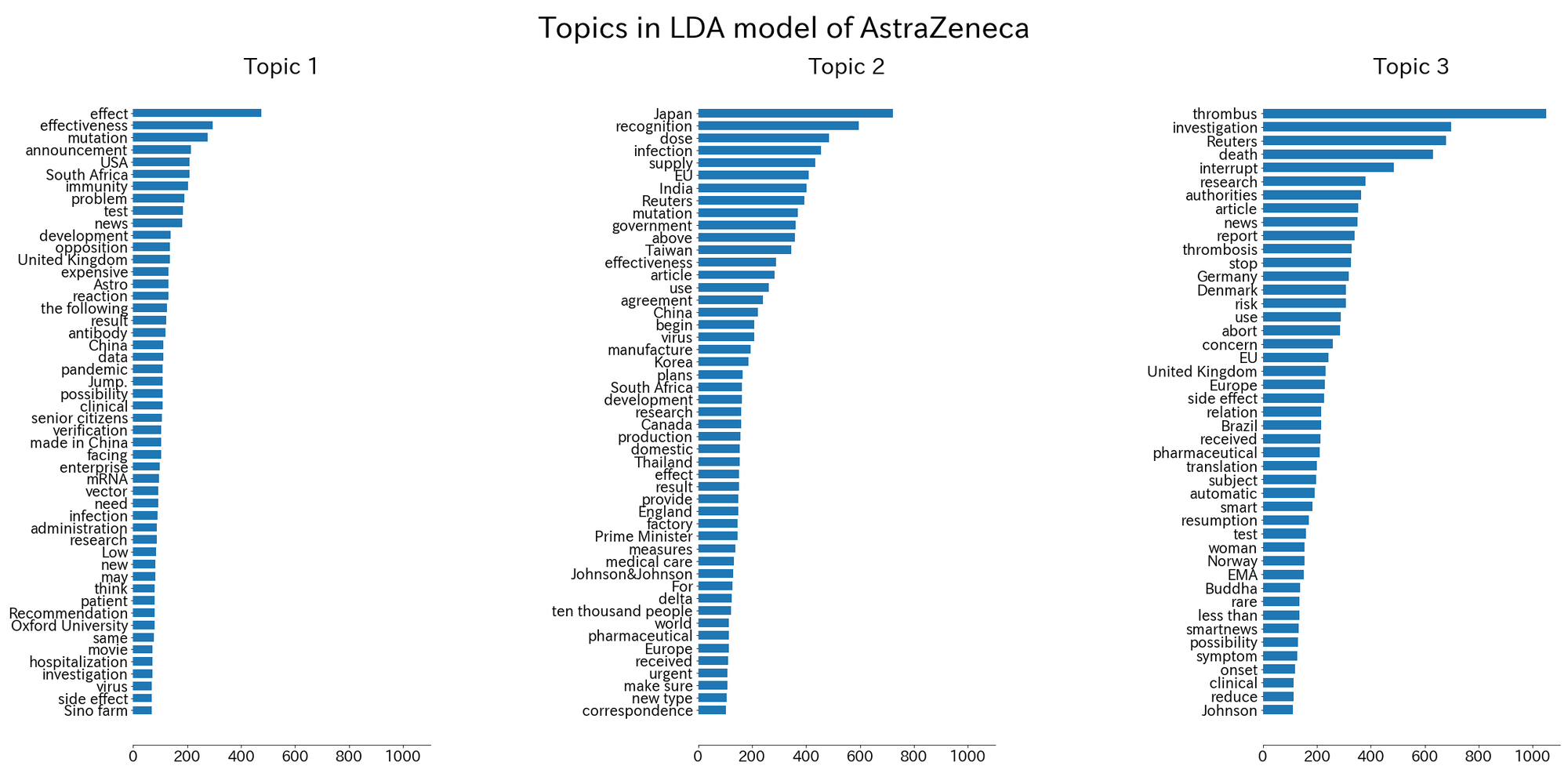

Figure 17. Word cloud of three LDA topics for tweets related to AstraZeneca vaccine (English version).

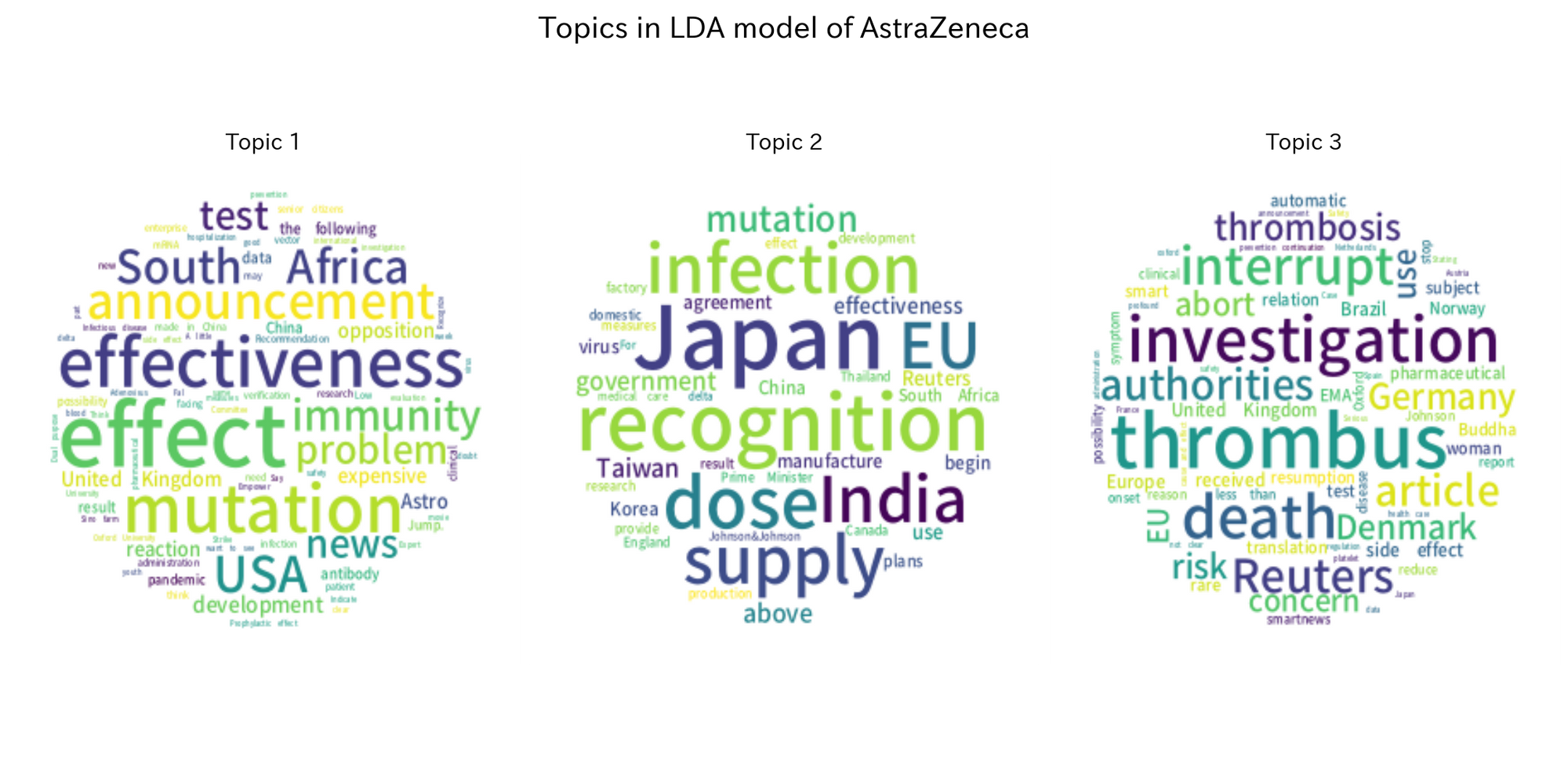

Figure 18. Top-50 words of three LDA topics for tweets related to Moderna vaccine (English version).

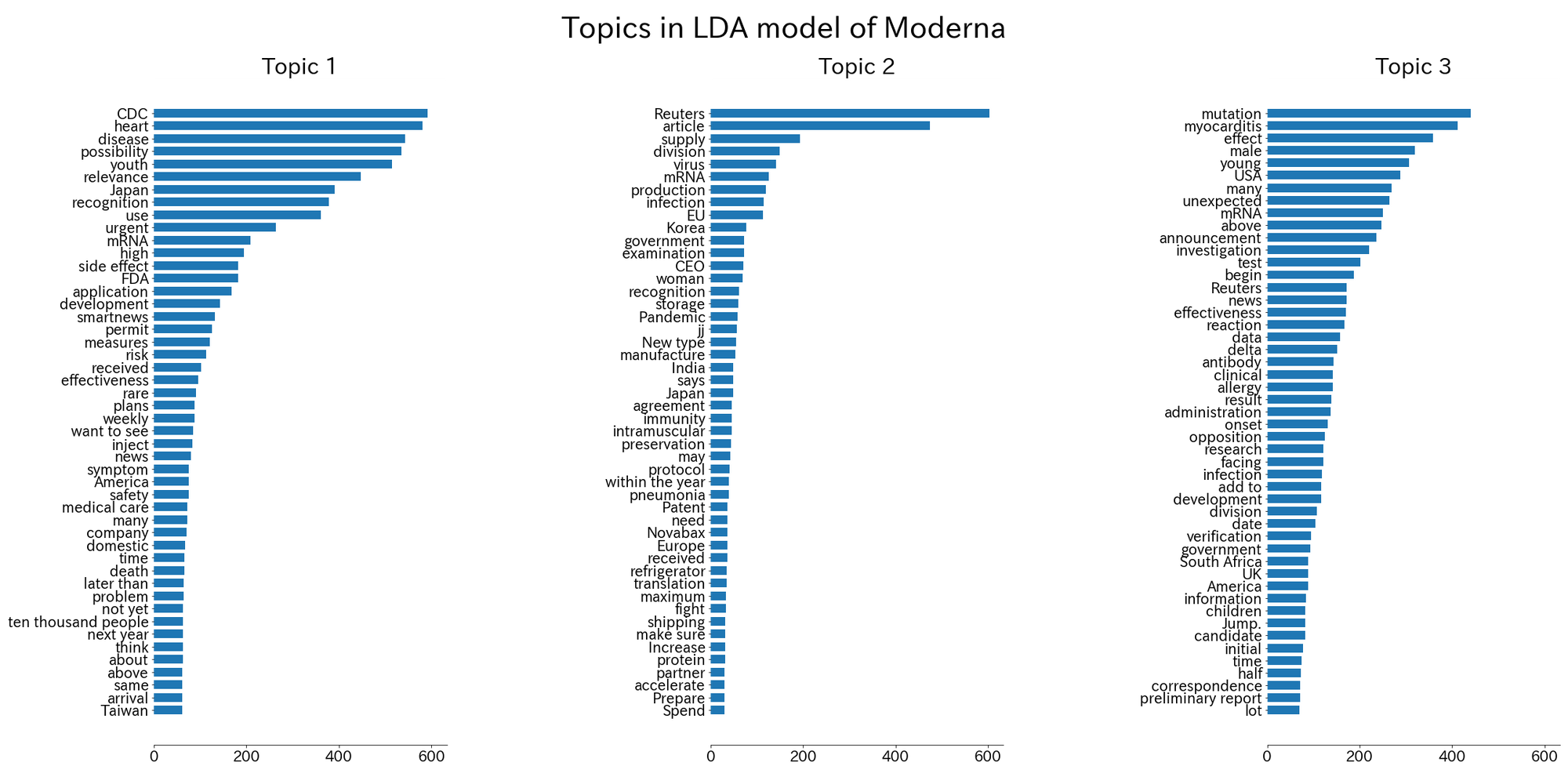

Figure 19. Word cloud of three LDA topics for tweets related to Moderna vaccine (English version).

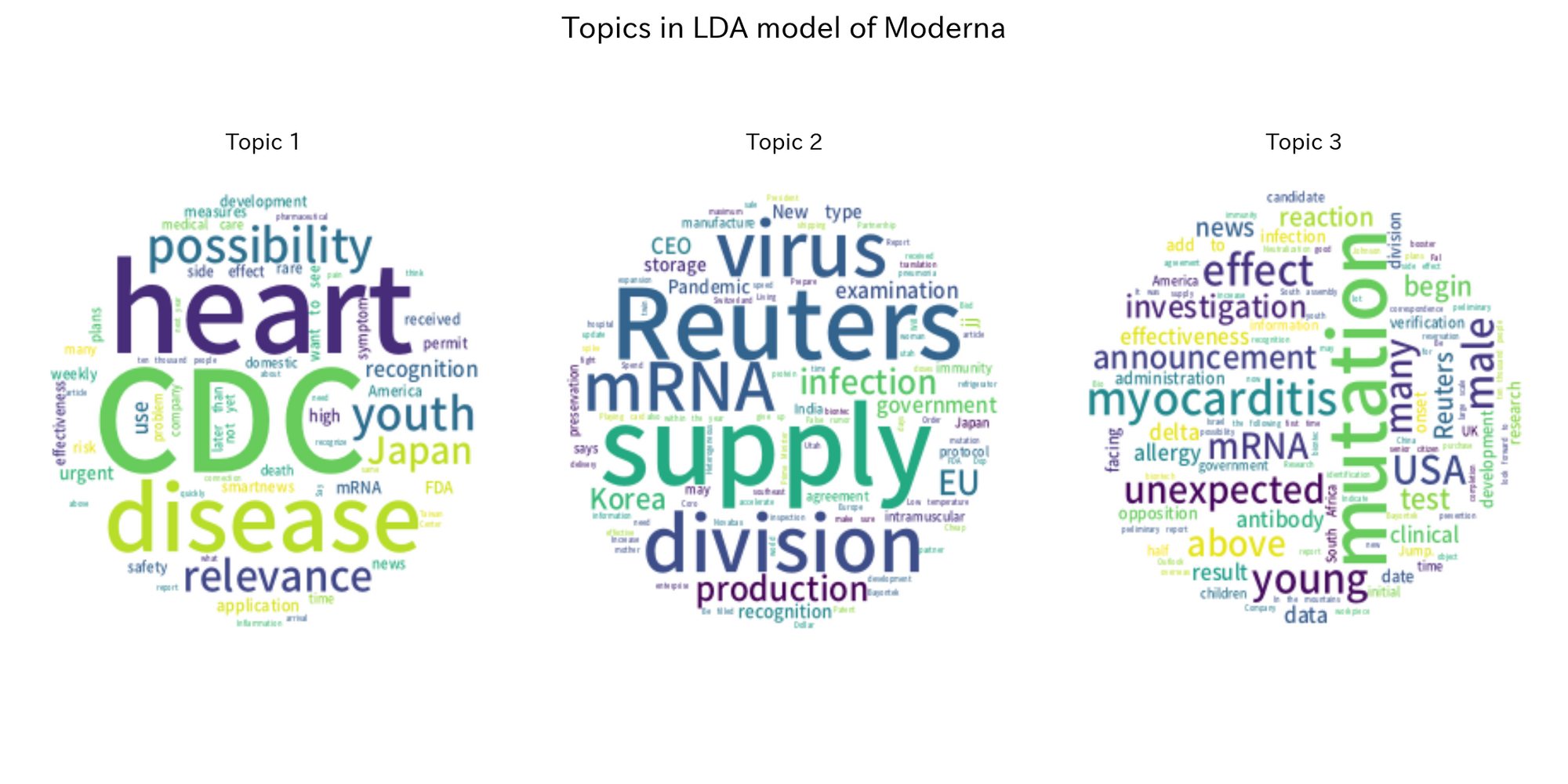
